# Dynamic associations between tau aggregation, atrophy, and cognitive decline in Alzheimer’s disease

**DOI:** 10.1101/2024.07.20.24310441

**Authors:** Ellen Hanna Singleton, Niklas Mattsson-Carlgren, Alexa Pichet Binette, Erik Stomrud, Olof Strandberg, Sebastian Palmqvist, Rik Ossenkoppele, Oskar Hansson

**Affiliations:** Clinical Memory Research Unit, Department of Clinical Sciences Malmö, Lund University, Sweden; Department of Neurology, Skåne University Hospital, Lund University, Lund, Sweden; Wallenberg Center for Molecular Medicine, Lund University, Lund, Sweden; Memory Clinic, Skåne University Hospital, Malmö, Sweden; Alzheimer Center Amsterdam, Department of Neurology, Amsterdam Neuroscience, Vrije Universiteit Amsterdam, Amsterdam UMC, Amsterdam, the Netherlands

**Keywords:** “Alzheimer’s disease”, “Tau PET”, “MRI”, “Cognition”, “Longitudinal”, “Clinical trials”

## Abstract

Tau aggregation measured with PET and neurodegeneration measured with MRI are closely associated with cognitive decline in Alzheimer’s disease, but the independent contributions of baseline and longitudinal measures of these imaging markers to cognitive decline remain unclear. Here, we tested a) independent associations between baseline and rate of change in tau-PET or MRI and cognitive decline, b) relative contributions of baseline and rate of change in tau-PET or MRI vs cognitive decline and c) effects of simulated treatment-induced reductions of tau aggregation on cognitive trajectories.

We included *n*=761 amyloid-positive individuals from the Swedish BioFINDER-2 study (*n*=253 cognitively unimpaired; *n*=508 cognitively impaired), with [^18^F]RO948-tau-PET, 3T structural MRI and cognition, including MMSE and a cognitive composite, of whom *n*=322 participants had longitudinal imaging data (*n*=120 cognitively unimpaired; *n*=202 cognitively impaired). Tau-PET SUVR and cortical thickness measures were quantified in entorhinal, amygdala (tau-PET only), inferior middle temporal, neocortical regions and hippocampal volume (MRI only). Associations between imaging measures and cognitive decline were assessed using linear mixed models with random intercepts and slopes or linear regressions, adjusting for age, sex, education, diagnosis and other imaging modality.

Baseline and longitudinal tau-PET showed stronger associations with cognitive decline than MRI, with the strongest effects for inferior-middle-temporal and neocortical regions for baseline tau-PET (MMSE: inferior-middle-temporal: *β*=-0.18±0.01, *p*<0.001; neocortical: *β*=-0.17±0.01, *p*<0.001; cognitive-composite: inferior-middle-temporal: *β*=-0.22±0.02, *p*<0.001; neocortical: *β*=-0.25±0.02, *p*<0.001), and neocortical composite for longitudinal tau-PET (MMSE: *β*=-0.56±0.04, *p*<0.001; and cognitive-composite: *β*=-0.62±0.05, *p*<0.001). When correcting for the other imaging modality, associations for tau-PET showed smaller reductions than MRI at baseline (14.9% for tau-PET vs 46.7% for MRI). Serial mediation models showed that baseline tau-PET explained the largest proportion of cognitive decline (54.0-94.0%), with modest mediation effects for longitudinal tau-PET or MRI pathways (2.0-15.0%). Simulated treatment-induced partial reduction of tau aggregation showed marginal effects on cognitive trajectories (MMSE: placebo vs 70% slope reduction: *Δ_rate-of-change_*=0.35[0.18-0.53], *p*<0.001, placebo vs 50%: *Δ_rate-of-change_*=0.25[0.08-0.43], *p*<0.001, cognitive-composite: placebo vs 70%: *Δ_rate-of-change_*=0.10[0.04-0.16], *p*<0.001) compared to halting tau progression (MMSE: placebo vs no-progression: *Δ_rate-of-change_*=1.03[0.85-1.22], *p*<0.001, cognitive-composite: placebo vs no-progression: *Δ_rate-of-change_*=0.33[0.27-0.39], *p*<0.001).

In conclusion, our results demonstrated the greater utility of tau-PET than MRI as a prognostic and disease monitoring marker, across the clinical spectrum of Alzheimer’s disease. In addition, baseline tau load showed stronger associations with future cognitive decline than tau progression. Furthermore, simulated reductions of tau progression showed limited effects on cognitive trajectories compared to halting tau progression and may point towards the need for early tau interventions.

## Introduction

*In-vivo* imaging measures of tau pathology and neurodegeneration, i.e. tau PET and MRI, have shown strong associations with cognitive decline in Alzheimer’s disease.^1,2^ Compared with MRI, tau PET showed higher accuracy for distinguishing Alzheimer’s disease from other neurodegenerative diseases^3,4^ and stronger predictive power for cross-sectional cognitive performance.^5–7^ In addition, tau PET has shown strong prognostic value in Alzheimer’s disease,^7–12^ even in early stages when no overt symptoms are present.^13^ While stronger independent associations with longitudinal cognition have been found for neurodegeneration in preclinical Alzheimer’s disease,^14^ superior associations for tau PET have been observed consistently in preclinical^9^ and general Alzheimer’s disease.^10,15,16^ However, large-scale head-to-head comparisons of *independent* effects of tau PET vs MRI on cognitive decline across clinical stages of Alzheimer’s disease are relatively lacking. Insights into these associations may provide guidance to select optimal markers to prognosticate Alzheimer’s disease, as well as to track disease progression, which is highly relevant for clinical trial design, both in terms of inclusion criteria^17^ and outcome measures.^18^

According to the temporal sequence of biomarker progression in Alzheimer’s disease,^19^ tau pathology is suggested to precede atrophy,^20^ and to drive cognitive decline.^15^ However, studies showing partial mediation effects of atrophy on the relationship between cross-sectional tau pathology and cognition, suggested alternative tau-mediated pathways to cognitive decline independent from neuronal loss.^5,6^ While baseline tau pathology and atrophy have shown complementary contributions to cognitive decline,^15,21^ the complex interplay between baseline and changes in tau pathology and neurodegeneration remains largely unknown. Mapping these complex interactions, similar to prior work assessing contributions of baseline and longitudinal components of amyloid-beta (Aβ) and tau pathology on cognitive decline in preclinical Alzheimer’s disease,^22^ may greatly enhance our understanding of specific determinants of cognitive decline in Alzheimer’s disease. In addition, these insights could inform therapeutic trials on the most optimal screening and disease progression markers.^23^

Therapeutic trials targeting tau pathology have shown relative inefficacy using tau antibodies aimed at inhibiting tau protein aggregation, which has led to the general consensus that that targeting tau aggregation may not be early or sufficient enough to prevent transsynaptic spread and cognitive benefits.^24^ Simulations of clinical trials in Alzheimer’s disease have been proven feasible based on real world data from large clinical research cohorts.^25^ Better insights into the impact of altering tau aggregation on cognitive trajectories may inform clinical trials on the effectiveness of reducing tau aggregation vs targeting accumulated tau load. With the recent developments in terms approval of anti-Aβ treatments^26^ and potential diversity in treatment effects among individuals with differing disease stages, combination therapies may be important in Alzheimer’s disease in the future,^23^ similar to oncological health care.^27^

Therefore, the aims of the current study were to investigate a) the independent associations between baseline and rate of change in tau PET or MRI and cognitive decline, b) the interplay between baseline and rate of change in tau PET or MRI and cognitive decline and, c) the effects of treatment-induced simulations of reductions tau aggregation on cognitive trajectories across the Alzheimer’s disease clinical continuum.

## Methods

### Participants

We included participants from the ongoing prospective Swedish BioFINDER-2 cohort (NCT03174938, http://www.biofinder.se/). All participants were recruited at Skåne University Hospital and the Hospital of Ängelholm, Sweden and the cohort covers the full spectrum of Alzheimer’s disease, ranging from Aβ-positive adults with intact cognition and no memory complaints (healthy controls) and subjective cognitive decline (SCD), to mild cognitive impairment (MCI) and Alzheimer’s disease dementia. The former two were labeled as cognitively unimpaired (CU) and the latter two as cognitively impaired (CI). The main inclusion criteria, as described previously,^28^ were age >40 years, fluency in Swedish language, and diagnoses were assigned based on Mini Mental State Examination (MMSE) scores as well as clinical evaluations including neuropsychological testing. Exclusion criteria were having significant unstable systemic illness, significant active alcohol or substance misuse, or refusing lumbar puncture or neuroimaging. MCI diagnosis was established if participants performed below 1.5 standard deviation from controls on at least one cognitive domain from an extensive neuropsychological battery examining verbal fluency, episodic memory, visuospatial ability, and attention/executive domains.^29^ Alzheimer’s disease dementia diagnosis was determined using criteria for dementia due to Alzheimer’s disease from the Diagnostic and Statistical Manual of Mental Disorders Fifth Edition and if positive on Aβ biomarkers based on the NIA-AA criteria for Alzheimer’s disease.^19^ All included participants were Aβ-positive based on CSF A*β*42/40 ratios, with a pre-established cutoff of 0.08 using the Elecsys immunoassays (Roche Diagnostics).^28^ Participants were included if they had at least one visit with tau PET, MRI and cognivite data. This led to the inclusion of n=253 CU Aβ-positive (n=125 cognitively normal controls and n=128 individuals with SCD) and n=508 CI Aβ-positive individuals (n=237 with MCI and n=271 with dementia). All data for the current study was acquired between April 2017 and October 2023.

### Standard protocol approvals, registrations and patient consents

All participants gave written informed consent. Ethical approval was given by the Regional Ethical Committee in Lund, Sweden. Approval for PET imaging was obtained from the Swedish Medical Products Agency and the local Radiation Safety Committee at Skåne University Hospital, Sweden.

### Image acquisition and processing of tau PET and MRI

MRI was performed using a Siemens 3T MAGNETOM Prisma scanner (Siemens Medical Solutions). Structural T1-weighted MRI images were acquired from a magnetization-prepared rapid gradient echo (MPRAGE) sequence with 1 mm isotropic voxels. PET images were acquired on digital GE Discovery MI scanners. For tau PET, acquisition was done 70–90 min post injection of ∼370 MBq [^18^F]RO948. Images were processed according to our pipeline described previously.^3^ Briefly, PET images were attenuation corrected, motion corrected, summed and registered to the closest T1-weighted MRI processed through the longitudinal pipeline of FreeSurfer version 6.0. Standardized uptake value ratio (SUVR) images were created using the inferior cerebellar gray matter as the reference region.^30^

### Regional tau PET and MRI

All PET and MR images were parcellated based on the Desikan-Killiany atlas,^31^ using FreeSurfer (FreeSurfer version 6.0) image analysis pipelines (surfer.nmr.mgh.harvard.edu). For tau PET, we computed SUVRs for entorhinal (weighted average of bilateral entorhinal cortices), amygdala (weighted average of bilateral amygdala volumes), inferior middle temporal (a weighted average of bilateral middle temporal and inferior temporal gyri) and neocortical (weighted average of all neocortical regions) regions of interest. The entorhinal, amygdala and inferior middle temporal regions were based on a well-established progression of tau pathology from the medial temporal lobe into the lateral temporal cortex^32,33^ as previously described.^13^ For MRI, cortical thickness was quantified in entorhinal, inferior middle temporal and neocortical regions using the same FreeSurfer subregions as for tau PET. In addition, hippocampal volume was calculated based on a weighted average of bilateral hippocampal volumes, corrected for intracranial volume.

### Cognition

For cognition, both the MMSE and the modified Preclinical Alzheimer’s Cognitive Composite- 5 (mPACC5) were used. A cognitive composite score analogous to the original PACC5 was calculated as a measure to capture early cognitive decline.^34,35^ The tests included were the MMSE, Alzheimer’s Disease Assessment Scale-Cognitive Subscale (ADAS-Cog) delayed recall, Trail-Making Test version A (TMT-A) and Category Fluency of animals. The original PACC5 includes two measures of memory recall; Logical Memory and the Free and Cued Selective Reminding Test. Given that in our cohort only one memory score was available, ADAS-cog delayed recall was assigned a double weight to maintain the same proportion of memory as in the original composite score.^36,37^ Further, the Digit Symbol Substitution Test used in the original PACC5 was replaced by TMT-A (to avoid missing values, as not all patients with MCI and dementia completed the Symbol Digit Modalities Test, as described previously^37^). All tests were z-scored based on the mean and standard deviation of CU Aβ- negative participants over 50 years old, and then averaged to generate cognitive composite scores.

### Statistical analysis

Demographic differences were assessed with two sample t-tests or chi-square tests where appropriate. We tested associations between baseline tau PET or MRI and longitudinal cognition using linear mixed models fitted with random intercepts and random slopes, adjusting for time, age, sex, education, and diagnosis (CU vs CI). The models included interactions for time with all other predictors.

Next, we tested associations between longitudinal tau PET or MRI (rate of change) and cognitive decline (rate of change) using a two-step approach. We first generated subject-level rates of change in tau PET, MRI and cognition, using linear mixed effect models with random intercepts and slopes for each brain region or cognitive test separately. These models used tau PET SUVR, MRI or cognition as the dependent variables and time (from the baseline scan or test date) as the independent variable. This yielded subject-level rates of change, for each imaging or cognitive measure, which were then used in subsequent analyses. We then tested associations between slopes of tau PET or MRI as predictors and slopes of cognition as outcomes (using the subject-level rates calculated before) using linear regression models, adjusting for age, sex, education and diagnosis (CU vs CI).

For models using baseline values or slopes of tau PET and MRI as predictors, models were fitted with and without correction for the other imaging modality to assess the independent effects of each modality on longitudinal cognition. To test significant differences between imaging predictors, models in the previous step were first bootstrapped (using n=1000 iterations) and subsequently compared using t-tests. Models were applied to the overall cohort and repeated in CU and CI groups separately in post-hoc analyses. In sensitivity analyses, models were repeated while adjusting for baseline values of the same modality and the average percent effect change was calculated for both modalities.

Next, to test relative contributions of baseline and longitudinal tau PET or MRI to longitudinal cognition, serial mediation models were performed. Models were constructed for the regions showing highest associations with longitudinal cognition in the previous steps, i.e. inferior middle temporal and neocortical regions. Models were constructed assessing the mediating effect of longitudinal tau PET or MRI on the associations between baseline tau PET and longitudinal cognition, adjusting for age, sex, education and diagnostic group (CU vs CI) and bootstrapping with n=5000 iterations.

To characterize to what degree reductions of tau aggregation impact the degree of cognitive decline we simulated effects on longitudinal cognition according to different interventions on tau PET slopes. We tested hypothetical reductions of tau PET slopes by 30%, 50% or 70%, similar to previously reported procedures.^38^ We then compared predicted cognitive slopes when reducing (i.e. simulated “treatment” arms) versus when not reducing tau PET slopes (i.e. simulated “placebo” group). In addition, we simulated an arm with no progression of tau PET signal using standard deviations from the original models to preserve variability (i.e. simulated “no progression” arm). Differences between treatment arms were assessed using ANOVAs with Bonferroni correction. All analyses were performed in R version 4.0.5. The main packages used were lme4 v1.1-30 for linear mixed effect models, lavaan v0.6- 13 for mediation analyses, predict v3.1-2 for simulation models and ggplot2 v3.3.6 for creating plots.

## Results

### Participants

We included *n*=253 CU Aβ-positive (Aβ+, *n*=125 cognitively normal controls and *n*=128 individuals with SCD) and *n*=508 CI Aβ-positive individuals (Aβ+, *n*=237 with MCI and *n*=271 with dementia). Demographic characteristics are presented in **Table 1**. The average follow-up time was 2.5±1.2 years in the overall cohort, 3.1±1.2 years in CU A+ and 2.3±1.2 years in CI Aβ+ individuals. The mean age was 73.0±7.9, 52% were female and the mean years of education was 13.0±4.1. At baseline the CI Aβ+ group had lower MMSE and mPACC5 scores, higher tau PET load, and lower cortical thickness and hippocampal volume (all *p*<0.05, **Table 1**). A subset of the cohort (*n*=120 CU Aβ+ and *n*=202 CI Aβ+ individuals) also had longitudinal imaging data available, with similar basic demographic characteristics as the overall sample (**Supplementary Table 1**). Imaging and cognitive slopes of this sample are shown in **Supplementary Table 1**.

**Table 1.** Demographic and clinical characteristics.

| | CU Aβ+ | CI Aβ+ | Overall | p-value |
| --- | --- | --- | --- | --- |
| *n* | 253 | 508 | 761 |  |
| Age, years | 73.0 (± 8.9) | 73.0 (± 7.3) | 73.0 (± 7.9) | 0.335 |
| Female, *n* (%) | 134 (53.0) | 265 (52.0) | 399 (52.0) | 0.896 |
| Education, years | 13.0 (± 3.7) | 12.0 (± 4.3) | 13.0 (± 4.1) | 0.435 |
| Follow-up time, years | 3.1 (± 1.2) | 2.3 (± 1.2) | 2.5 (± 1.2) | <0.001 |
| MMSE | 29.0 (± 1.3) | 24.0 (± 4.4) | 25.0 (± 4.4) | <0.001 |
| mPACC5 | -0.3 (± 0.8) | -2.9 (± 1.7) | -2.0 (± 1.9) | <0.001 |
| Tau PET SUVR |  |  |  |  |
| Entorhinal | 1.4 (± 0.3) | 1.8 (± 0.4) | 1.7 (± 0.5) | <0.001 |
| Amygdala | 1.1 (± 0.3) | 1.6 (± 0.5) | 1.4 (± 0.5) | <0.001 |
| Inferior middle temporal | 1.3 (± 0.3) | 1.9 (± 0.7) | 1.7 (± 0.7) | <0.001 |
| Neocortical | 1.1 (± 0.1) | 1.4 (± 0.4) | 1.3 (± 0.4) | <0.001 |
| MRI cortical thickness, mm<sup>2</sup> |  |  |  |  |
| Entorhinal | 3.1 (± 0.3) | 2.8 (± 0.4) | 2.9 (± 0.4) | <0.001 |
| Hippocampal volume | 0.0023 (± 0.0003) | 0.0020 (± 0.0003) | 0.0021 (± 0.0003) | <0.001 |
| Inferior middle temporal | 2.5 (± 0.1) | 2.4 (± 0.1) | 2.4 (± 0.1) | <0.001 |
| Neocortical | 2.3 (± 0.1) | 2.2 (± 0.1) | 2.3 (± 0.1) | <0.001 |

Data are shown as mean (standard deviation) unless otherwise specified. *P*-values for CU Aβ + versus CI Aβ +, using chi-square test for categorical variables and ANOVA's for continuous variables. Aβ +=amyloid
positive, CU=cognitively unimpaired, CI=cognitively impaired, MMSE=mini mental state evaluation, mPACC5=modified preclinical Alzheimer's cognitive composite, SUVR=standardized uptake value ratio.

### Associations between baseline tau PET or MRI and cognitive decline

When assessing the associations between baseline tau PET or MRI and cognitive decline, the strongest predictors of longitudinal MMSE were tau PET in the inferior middle temporal (*β*=- 0.18±0.013, *p*<0.001) and neocortical regions (*β*=-0.17±0.01, *p*<0.001). The MMSE score thus decreased 0.18 and 0.17 standard deviations respectively per year for every standard deviation of increase in tau PET SUVR at baseline. The next best predictor was cortical thickness in inferior middle temporal regions (*β*=-0.11±0.01, *p*<0.001). For mPACC5, the strongest predictors were tau PET in the inferior middle temporal region (*β*=-0.22±0.02, p<0.001), the neocortical region (*β*=-0.25±0.02, p<0.001), and the entorhinal region (*β*=-0.11±0.02, p<0.001; **Fig. 1** and **Supplementary Table 2**).

**Figure 1.**
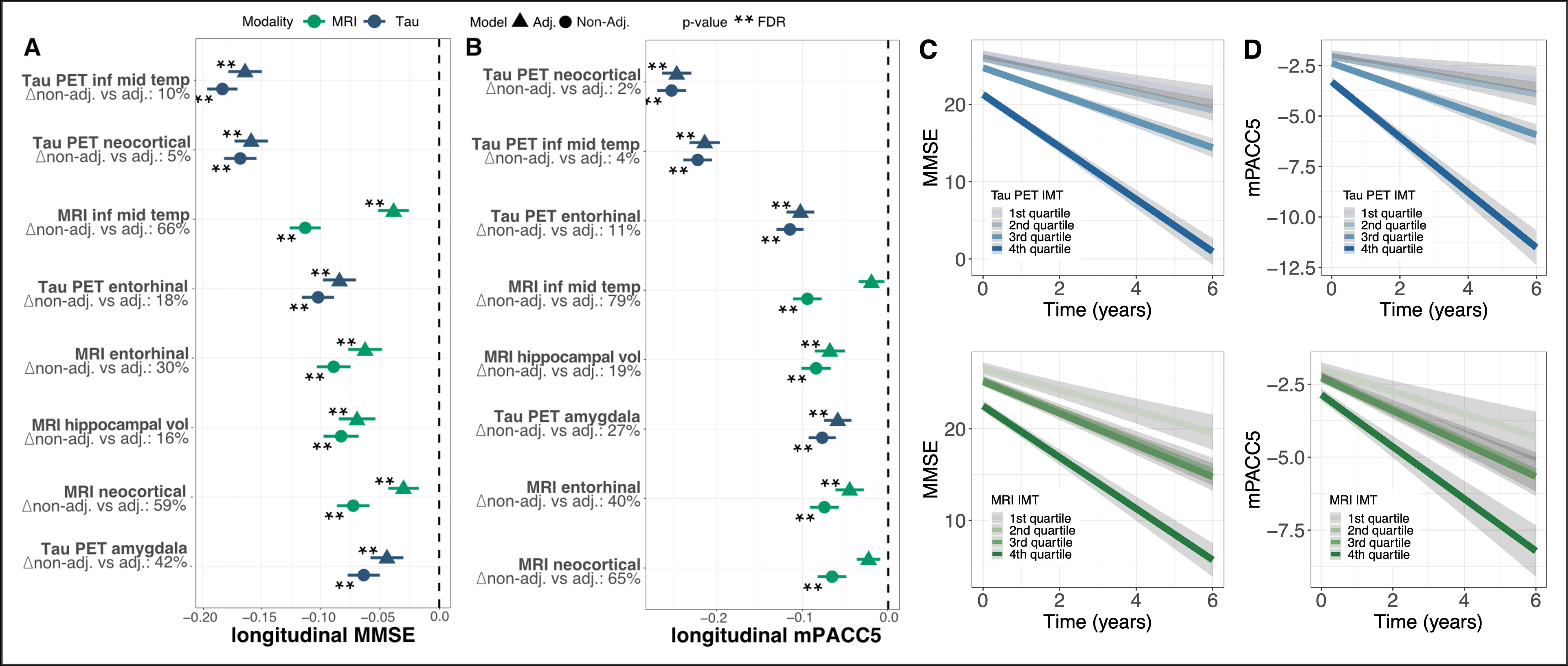
Associations between baseline tau PET or MRI and longitudinal cognitive decline in CU&CI Aβ+ individuals. **(A)** shows associations between baseline tau PET and MRI and MMSE in the whole cohort (CU Aβ+ and CI Aβ+ individuals combined) while adjusting for age, sex, education and cognitive status (CU vs CI). **(B)** shows these associations for the mPACC5. Standardized coefficients for tau PET are depicted in blue, and standardized coefficients for MRI are depicted in green. Circles represent models without adjustment for the other modality, while triangles represent models with adjustment for the other imaging modality. **=significant after FDR correction, *=significant at *p*=0.05. **(C)** and **(D)** show changes in MMSE and mPACC5 by quartiles of neocortical tau PET (blue) or MRI (green), adjusted for age, sex and education.

For the MMSE, the effects in most regions were significantly different from the next best region, except for MRI entorhinal vs MRI hippocampal volume (*Δβ*=-0.013[-0.028, 0.003], **Supplementary Table 3**). For the mPACC5 all effects were significantly different from the next predictor, except for MRI inferior middle temporal vs MRI hippocampal volume (*Δβ*=-0.004[-0.025,0.017], **Supplementary Table 3**) and MRI entorhinal vs MRI neocortical (*Δβ*=0.014[-0.005,0.034], **Supplementary Table 3**). Similar results were obtained when repeating analyses for CU Aβ+ and CI Aβ+ groups separately (**Supplementary Fig. 2-3** and **Supplementary Tables 4-5**).

We next adjusted the tau PET models for MRI in the same region, and vice versa. This reduced the effects markedly for some of the MRI measures (especially inferior middle temporal and neocortical regions), while tau PET effects were generally more robust (mean reduction of effect: 14.9% for tau PET vs 46.7% for MRI; **Fig. 1**). After this adjustment, tau PET measures were the strongest predictors for both MMSE (inferior middle temporal: *β*=- 0.16±0.01, *p*<0.001, reduction: 10.5% when adjusting for MRI; neocortical: *β*=-0.16±0.01, *p*<0.001, reduction: 5.5%; entorhinal: *β*=-0.10±0.01, *p*<0.001, reduction: 17.7%) and mPACC5 (neocortical: *β*=-0.25±0.02, *p*<0.001, reduction: 2.4%; inferior middle temporal: *β*=-0.21±0.02, *p*<0.001, reduction: 3.6%; entorhinal: *β*=-0.10±0.02, *p*<0.001, reduction: 10.7%; **Fig. 1, Supplementary Fig. 1** and **Supplementary Table 2**).

### Associations between longitudinal tau PET or MRI and cognitive decline

We next tested associations between regional longitudinal tau PET or MRI (rate of change) and cognitive decline (rate of change). Change in tau PET levels in the neocortical region showed the strongest associations with change in MMSE (*β*=-0.56±0.04, *p*<0.001) and change in mPACC5 (*β*=-0.62±0.05, *p*<0.001), followed by inferior middle temporal cortical thickness (MMSE: *β*=-0.55±0.04, *p*<0.001, mPACC5: *β*=-0.48±0.05, *p*<0.001) and inferior middle temporal tau PET (MMSE: *β*=-0.48±0.05, mPACC5: *β*=-0.47±0.05, *p*<0.001; **Fig. 2** and **Supplementary Table 6**). For both tests, all predictors were significantly different from the next best predictor e.g., first vs second rank, and second vs third rank, except for MRI neocortical vs MRI hippocampal volume for MMSE (*Δβ*=-0.025[-0.188-0.133]) for MRI neocortical vs MRI hippocampal volume for mPACC5 (*Δβ*=0.084[-0.096-0.275], **Supplementary Table 7**).

**Figure 2.**
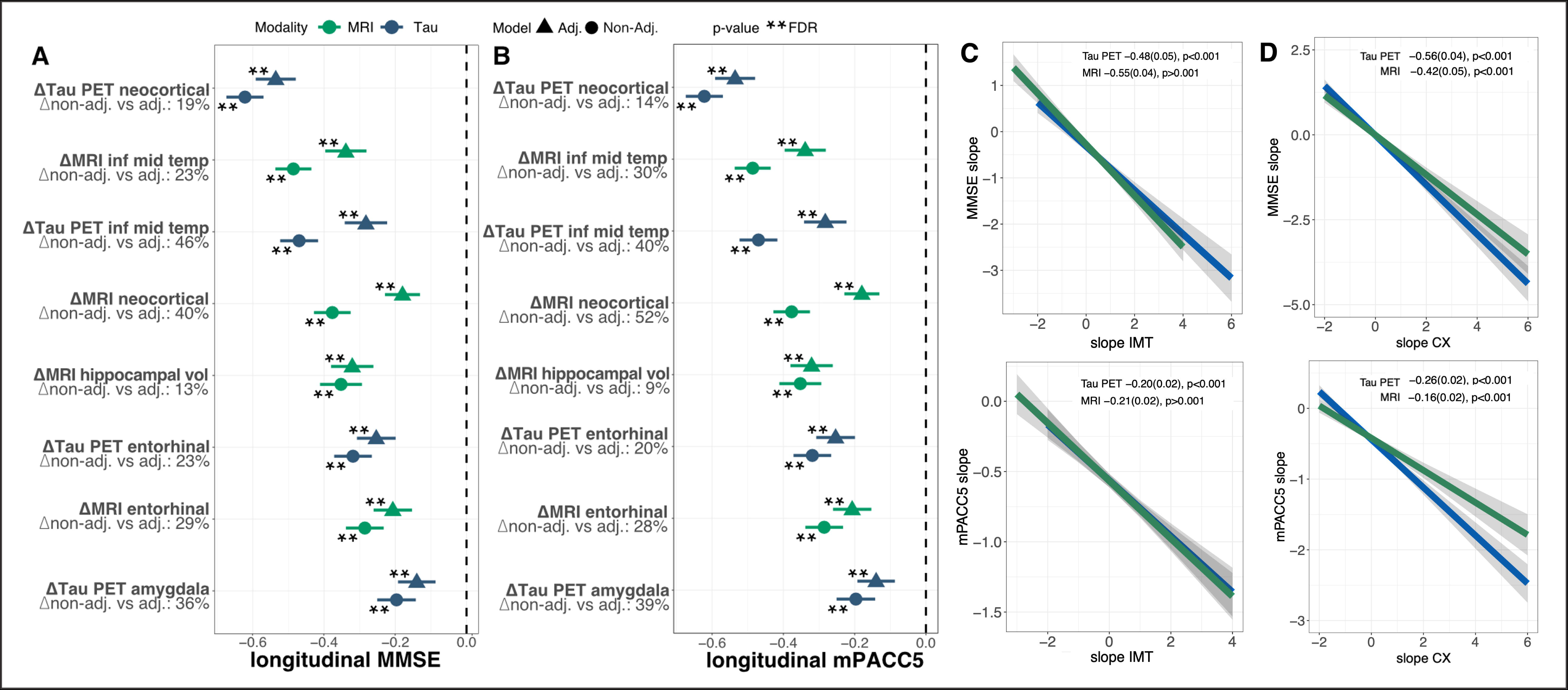
Associations between rate of change in tau PET or MRI and rate of change in cognition in CU&CI Aβ+ individuals. **(A)** shows associations between slope in tau PET and MRI and slope in MMSE in the whole cohort (CU Aβ+ and CI Aβ+ individuals combined) while adjusting for age, sex, education and cognitive status (CU vs CI), while **(B)** shows these associations for the mPACC5. The slopes were obtained from linear mixed models with random slopes and random intercepts, assessing either tau∼time, mri∼time, or cognition∼time, yielding average rate of change metrics per individual. These were used as input for the linear regressions depicted in these panels. **(C)** and **(D)** show associations between rate of change in MMSE and mPACC5 and rate of change in tau PET or MRI in regions of interest. **=significant after FDR correction, *=significant at p=0.05.

When adjusting these models for rate of change in the other modality, the strongest predictors remained similar for both MMSE (tau PET neocortical: *β*=-0.46±0.05, *p*<0.001, reduction: 18.6%; MRI inferior middle temporal: *β*=-0.42±0.05, *p*<0.001, reduction: 23.5%; tau PET inferior middle temporal: *β*=-0.26±0.05, *p*<0.001, reduction: 45.7%) and mPACC5 (tau PET neocortical: *β*=-0.54±0.06, *p*<0.001, reduction: 13.9%; MRI inferior middle temporal: *β*=-0.34±0.06, *p*<0.001, reduction: 30.3%; tau PET inferior middle temporal: *β*=-0.28±0.06, *p*<0.001, reduction: 39.9%; **Fig. 2** and **Supplementary Table 6**). Similar results were obtained when repeating the analyses for CU Aβ+ and CI Aβ+ individuals separately (**Supplementary Fig. 4-5** and **Supplementary Table 8**-**9**). Overall, rate of change measures of tau PET and MRI showed similar reductions of effects after correction for the other modality (reduction of effect: 29.6% for tau PET vs 28.1% for MRI). Next, these models were fit while also adjusting for baseline imaging levels (**Supplementary Fig. 6**, **Supplementary Table 10-12)**. This had more pronounced effects for tau PET, where the rate of change effects were generally strongly reduced, while effects for rate of change in MRI were less affected by adjustment for baseline MRI (**Supplementary Fig. 6**, **Supplementary Table 10-12**).

### Contributions of baseline vs change in tau PET or MRI to cognitive decline

Next, we modelled the relationship between baseline and longitudinal imaging measures and cognitive decline using serial mediation analyses. We used the regions with strongest associations with longitudinal cognition in previous steps, i.e., inferior middle temporal and neocortical regions. Baseline tau PET was used as the main predictor, while tau PET rate of change, and baseline and rate of change MRI measures were used as mediators in associations with cognitive decline (**Supplementary Fig. 3** and **Supplementary Table 13-14**).

Overall, the largest proportion of longitudinal cognition was explained directly by baseline tau PET, ranging from 82.0-90.0% of the total effect explained in inferior middle temporal and 54.0-94.0% in neocortical regions (MMSE, inferior middle temporal: *β*=-0.48[- 0.71, -0.26], *p*<0.001, neocortical, *β*=-0.26[-0.53, -0.01], *p*<0.05; mPACC5, inferior middle temporal: -0.47[-0.75, -0.16], *p*<0.001; neocortical: -0.47[-0.76,-0.08], *p*<0.01; **Fig. 3** and **Supplementary Table 13-14**). In the inferior middle temporal region, the effect was partially mediated through MRI rate of change (*indirect pathway 1*; MMSE: *β*=-0.07[-0.17,-0.008], *p*<0.05, proportion mediated: 0.14; mPACC5: *β*=-0.08[-0.19,-0.01], p<0.05, proportion mediated: 0.15) and baseline MRI and MRI rate of change (*indirect pathway 2*; MMSE: *β*=- 0.05[-0.09,-0.03], *p*<0.05, proportion mediated: 0.10; mPACC5: *β*=-0.040[-0.076,-0.017], *p*<0.05, proportion mediated: 0.08). In the neocortical region, the effect was partially mediated through tau PET rate of change and MRI rate of change (*indirect pathway 1*; MMSE: *β*=-0.05[- 0.12,-0.01], *p*<0.05, proportion mediated: 0.10; mPACC5: *β*=-0.05[-0.12,-0.01], *p*<0.05, proportion mediated: 0.09) and baseline MRI and MRI rate of change (*indirect pathway 2*; MMSE: *β*=-0.02[-0.05,-0.01], *p*<0.05, proportion mediated: 0.05; mPACC5: *β*=-0.01[-0.40,- 0.003], *p*<0.05, proportion mediated: 0.02).

**Figure 3.**
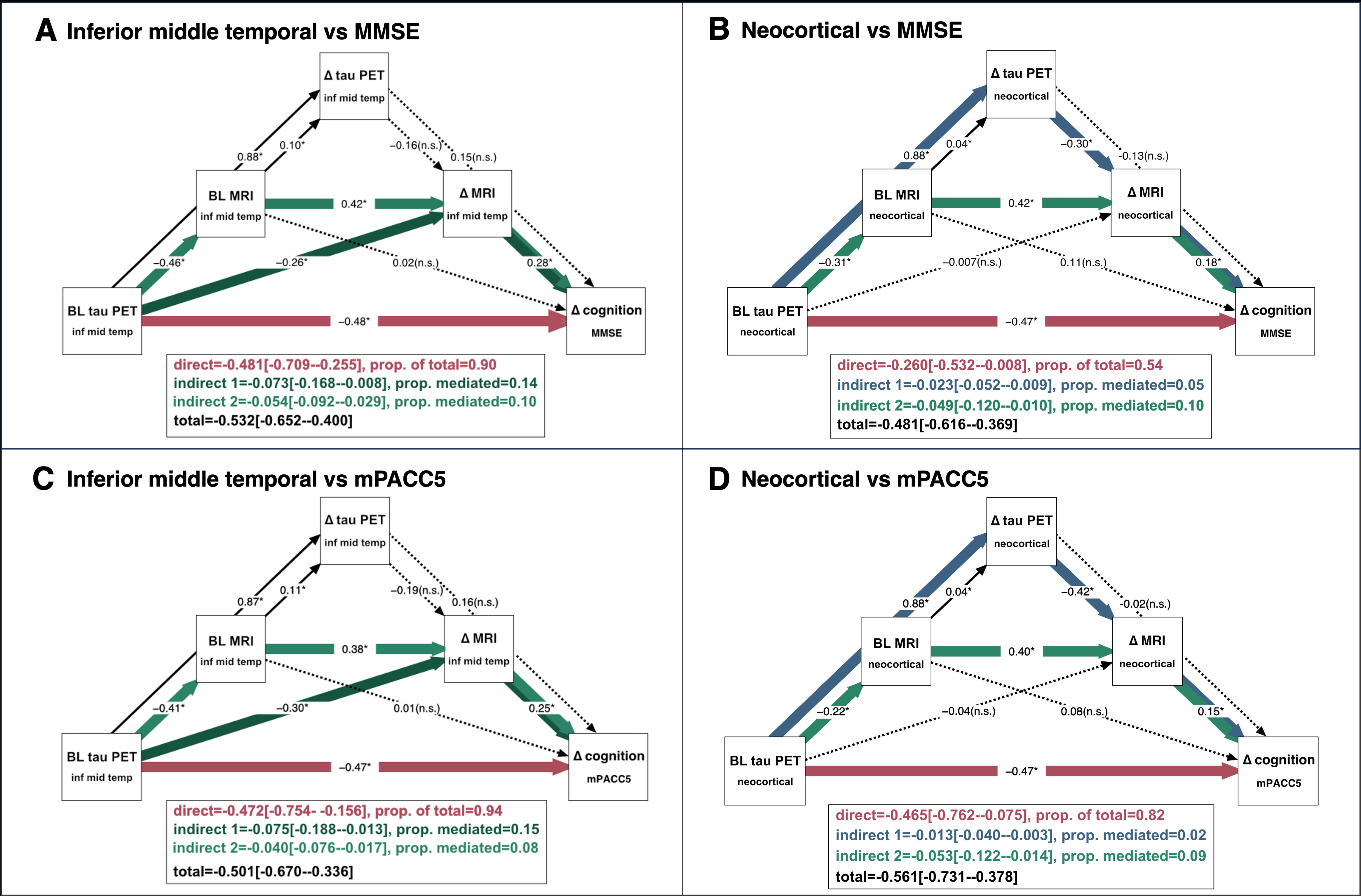
Contributions of baseline vs change in tau PET and MRI to cognitive decline in CU&CI Aβ+ individuals. **(A)** and **(B)** show associations between (rate of change in) tau PET, MRI and rate of change in MMSE in inferior middle temporal and neocortical regions respectively in the whole cohort (CU Aβ+ and CI Aβ+ individuals combined). **(C)** and **(D)** shows associations between (rate of change in) tau PET, MRI and rate of change in mPACC5 in inferior middle temporal and neocortical regions respectively in the whole cohort (CU Aβ+ and CI Aβ+ individuals combined). Analyses were adjusted for age, sex, education and cognitive status (CU vs CI), using n=5000 bootstrapping iterations. Solid lines indicate significant effects, while dotted lines indicate nonsignificant associations.

### Cognitive trajectories according to simulated reductions of tau aggregation

To assess the effect of interventions on tau aggregation on cognitive trajectories we simulated interventions on tau PET slopes and examined the effects on cognitive trajectories. We simulated reductions of 30%, 50% and 70% on tau PET slopes as well as a “no progression” treatment arm. Results for inferior middle temporal and neocortical regions are shown in **Fig. 4** and **Supplementary Table 15a-15b**. When assessing the neocortical region of interest, we observed marginal differences in cognitive slopes between placebo and treatment arms with tau PET slope reductions (MMSE, placebo:-1.3±1.0 vs 30%:-1.2±0.86, 50%:-1.1±0.76, 70%:- 0.95±0.68; mPACC5, placebo:-0.39±0.33 vs 30%:-0.35±0.28, 50%:-0.32±0.25, 70%:-0.29±0.23, **Fig. 4** and **Supplementary Table 15b**). Differences between treatment arms were observed in placebo vs 50% (*Δ_rate-of-change_*=0.25[0.08-0.43], *p*<0.001) and placebo vs 70% (*Δ_rate- of-change_*=0.35[0.18,0.53], *p*<0.001, Bonferroni corrected) for MMSE, and placebo vs 70% for mPACC5 (*Δ_rate-of-change_*=0.10[0.04,0.16], *p*<0.001, **Fig. 4** and **Supplementary Table 15b**). When assessing cognitive trajectories in placebo vs the “no progression” treatment arm we observed relatively larger differences (MMSE; placebo vs no progression: *Δ_rate-of- change_*=1.03[0.85-1.22], *p*<0.001, mPACC5; placebo vs no progression: *Δ_rate-of-change_*=0.33[0.27- 0.39], *p*<0.001, **Fig. 4** and **Supplementary Tables 15a-15b**) compared to placebo vs reduction treatment arms. Similar results were observed for the inferior middle temporal region of interest (**Fig. 4** and **Supplementary Table 15a**). When assessing CU Aβ+ and CI Aβ+ separately, no significant differences were observed between reduction treatment arms for CU Aβ+, while the “no progression” arm differed from placebo (*Δ_rate-of-change_*=1.03[0.90-1.16], *p*<0.05, **Supplementary Fig. 7** and **Supplementary Tables 17a-18b**). For CI Aβ+, all treatment arms differed from placebo, as well as 30% vs 70% reduction treatment arms (all *p*<0.05, Bonferroni corrected, **Supplementary Fig. 7** and **Supplementary Tables 17a-18b**).

**Figure 4.**
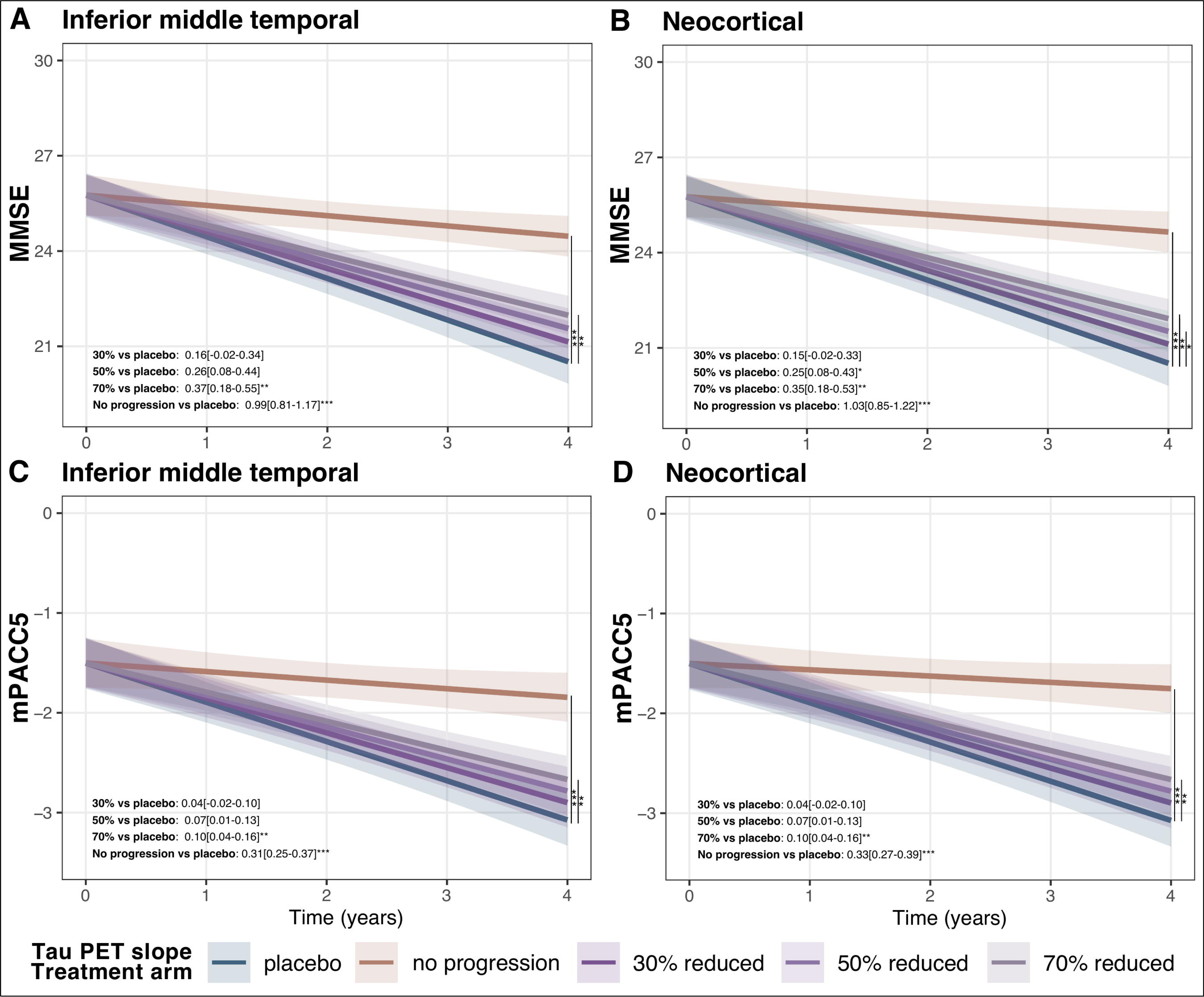
Cognitive trajectories according to simulated treatment-induced reductions of tau aggregation in CU&CI Aβ+ individuals. **(A)** and **(B)** show predicted MMSE values over time in inferior middle temporal and neocortical regions respectively in the whole cohort (CU Aβ+ and CI Aβ+ individuals combined) in different arms of tau PET slopes, i.e. placebo (no intervention) versus 30%, 50% and 70% reductions of tau PET slopes, while **(C)** and **(D)** show predicted mPACC5 values over time in inferior middle temporal and neocortical regions respectively in the whole cohort (CU Aβ+ and CI Aβ+ individuals combined).

## Discussion

In an Aβ+ cohort including cognitively unimpaired and impaired individuals, baseline and rate of change tau PET showed superior independent associations with cognitive decline, compared to MRI. In addition, baseline tau PET showed the largest direct contribution to cognitive decline and was only partially mediated by rate of change tau PET or MRI. Simulated treatment-induced reductions of tau PET aggregation showed more marginal attenuations of cognitive trajectories than halting tau progression. Taken together, this study demonstrated the utility of tau PET as a prognostic and disease monitoring marker relative to MRI and suggested baseline tau PET as a strong determinant of future cognitive decline in Alzheimer’s disease.

Regional tau PET markers showing the strongest associations with cognitive decline at baseline included inferior middle temporal and neocortical composite regions. While the entorhinal cortex^39^ or medial temporal cortex^40^ are suggested as early tau PET marker and inferior middle temporal as a later stage marker,^39^ the prognostic value of tau PET positivity in inferior middle temporal regions in early stages of Alzheimer’s disease has been demonstrated previously.^13^ When assessing tau PET rate of change as predictor, the neocortical composite showed strongest independent associations with cognitive decline. This is consistent with recent work in a smaller sample, showing strongest associations between neocortical tau PET and cognitive decline comparing tau pathology and neurodegeneration in Alzheimer’s disease.^15^ The substantial heterogeneity in tau pathology patterns and associated differential progression patterns in Alzheimer’s disease may favor the use of whole brain neocortical regions rather than specific one-size-fits-all regions of interest.^41^ Alternatively, the uptake in neocortical composites may reflect more advanced tau pathology associated with faster cognitive decline. While increased tau PET in either inferior middle temporal or neocortical composite regions may be efficient measures for baseline enrichment of individuals with high risk of cognitive decline due to Alzheimer’s disease, the uptake in a neocortical composite may be more informative as a disease monitoring marker, e.g., to track treatment effects in clinical trials.

Direct effects of tau pathology on baseline^5^ and longitudinal cognition^21^ have been observed previously in Alzheimer’s disease, which are independent of neurodegeneration. While the direct effects of *changes in* tau pathology on cognitive decline remain largely unknown, associations between longitudinal tau PET and cognitive decline were observed in a cognitively unimpaired population in the absence of associations between tau accumulation and structural changes.^42^ Our study represents one of the first large-scale cohorts to confirm superior independent associations between change in tau PET and cognitive decline compared to change in MRI. The mechanisms by which tau pathology may exert these direct effects, both at baseline and longitudinally, remain unclear. Tau pathology may impact neuronal function prior to neuronal loss by various mechanisms including early neurotoxicity of neurofibrillary tangle bearing neurons, subtle morphometric neuronal changes, synapse reductions, global network dysfunctions^5,43^ and microglial changes leading to chronic innate neuroinflammation^44,45^ or vascular mechanisms.^46,47^ Indeed, functional brain networks have been associated with both baseline and rate of change measures of tau PET previously.^43,48,49^ Alternatively, MRI measures may show considerable variability at baseline which do not depend on Alzheimer’s disease pathology, such as premorbid differences, non-pathological effects of aging on brain structure, and atrophy due to non- Alzheimer’s disease processes,^50–52^ impacting their specificity for detecting Alzheimer’s disease-related cognitive changes. Note that our mediation models showed that the effects of tau accumulation on cognitive decline are still to some degree dependent on neurodegeneration, supporting their complementary contributions.^12,15^

Baseline tau pathology was more strongly associated with cognitive decline than tau aggregation rates, when combined in serial mediation models. While similar associations between baseline and rate of change in tau PET and cognitive decline have been observed in a smaller Alzheimer’s disease cohort^53^ and stronger associations between change in tau pathology and cognitive decline than baseline tau pathology have been observed in preclinical Alzheimer’s disease,^22^ our study provides robust evidence for the importance of baseline vs longitudinal tau PET based on the systematic large-scale cohort and comprehensive modelling techniques across clinical stages. The relative importance of baseline tau PET may partly be due to the strong associations between tau aggregation rates and baseline tau load.^39,53^ Baseline tau PET load represents the cumulative accumulation of tau pathology over the course of many years, and it may be that the total amount of aggregated tau continuously exerts neurotoxic effects in the brain, which are not only due to newly formed tau aggregates (i.e. tau PET rate of change). Alternatively, test-retest reliability of tau PET may hamper the utility of longitudinal tau PET as a prognostic marker, which is higher for MRI and FDG-PET, where change metrics were more associated with cognitive decline.^54,55^ Future work should focus on head-to-head comparisons between tau PET and fluid markers more specific to tau pathology, such as MTBR-tau^56^ or p-tau205^57^ to assess the most potent and cost-effective disease progression markers in Alzheimer’s disease.

Simulated clinical trials with reductions of tau aggregation showed rather limited impact on cognitive trajectories in different treatment arms with reductions of tau aggregation compared to placebo, while halting tau progression showed larger effects on cognitive decline compared to placebo. Current therapeutic strategies targeting tau include strategies aimed at a) preventing translation of tau mRNA, b) altering tau post-translational modifications, or c) inhibiting spread of misfolded tau or inhibiting tau aggregation.^24^ The relative inefficacy of tau interventions using tau antibodies aimed at inhibiting tau protein aggregation, combined with the relative success of an antisense oligonucleotide targeting tau mRNA translation mechanisms^58,59^, suggests that targeting tau proteins may not be early or sufficient enough to prevent transsynaptic spread.^24^ Although speculative based on our simulated data, we carefully infer that these simulations suggest that complete removal of tau progression may be necessary to convey clear effects on cognitive trajectories. This may point to the need of early tau interventions, as has been suggested for anti-amyloid trials.^60^ However, this may be partly explained by the high dependence of tau PET progression on baseline tau PET load; it is therefore important to note that altering tau PET progression may still be effective in groups of patients with low baseline tau PET loads. Artificial intelligence solutions may be valuable tools in simulating effects of absolute tau load reductions on cognitive decline, simulating treatment effects at different levels of baseline tau load in the future. Improving clinical trial design for anti-tau therapies may eventually facilitate the field to move towards combination therapies in Alzheimer’s disease.^23^

A strength of this study is the standardized assessments of longitudinal multimodal imaging and cognition in a large sample across the entire Alzheimer’s disease clinical continuum. One limitation is that we only relied on structural MRI as a measure of neurodegeneration. However, we note that another study with FDG-PET showed similar results to ours, i.e., superiority of tau PET in associations with cognitive decline and only partial mediation of the relationship between tau PET and cognitive decline.^15^ Furthermore, the influence of other co-pathologies, e.g., TAR DNA-binding protein 43 (TDP-43) pathology contributing to cognitive decline cannot be ruled out. Future work may incorporate direct markers of co-pathologies into more comprehensive models, including CSF α-synuclein measures that have shown associations with longitudinal cognition,^29,61,62^ as well as potential novel plasma measures of TDP-43 pathology.^63^ In addition, due to large sample size requirements (sample sizes of *n*=200 or larger have been advocated for these models^64^), the serial mediation analyses were only performed in the combined cohort, and were not repeated within diagnostic subgroups. Based on the similar results obtained in the first steps of analyses between CU Aβ+ and CI Aβ+ individuals, disease-stage-specific effects in the serial mediation analysis were deemed marginal. Lastly, it should be noted that it is unclear how our simulation models translate to real world clinical trial data. It is furthermore possible that cognitive effects may take longer to appear than clinical trial durations, and there might still be effects of slowing tau aggregation which are difficult to predict from an observational study or data with heterogeneous baseline tau PET levels.

In conclusion, we observed a) greater independent associations between baseline and rate of change in tau PET and cognitive decline compared to baseline and rate of change in MRI b) greater contributions of baseline tau pathology vs tau aggregation over time to cognitive decline and c) marginal effects of treatment-induced reductions of tau aggregation on cognitive trajectories compared to halting tau progression in Alzheimer’s disease. This study demonstrated the utility of tau PET as an independent prognostic and disease monitoring marker throughout the clinical course of Alzheimer’s disease. In addition, it suggested baseline tau load as a strong determinant of future cognitive decline and may point towards the need for early interventions in therapeutic trials targeting tau pathology.

## Data availability

Anonymized data will be shared by request from a qualified academic investigator for the sole purpose of replicating procedures and results presented in the article and as long as data transfer is in agreement with EU legislation on the general data protection regulation and decisions by the Ethical Review Board of Sweden and Region Skåne, which should be regulated in a material transfer agreement.

## Acknowledgements

We would like to acknowledge all participants in the Swedish BioFINDER study for their participation.

## Abbreviations

Aβ: β amyloid
AD: Alzheimer’s disease
MCI: mild cognitive impairment
MMSE: mini mental state examination
mPACC5: modified Preclinical Alzheimer’s Cognitive Composite
SCD: subjective cognitive decline.

## Supplementary material

**Supplementary material** is available at *Brain* online.

