## Supplementary Material for "Dynamic associations between tau aggregation, atrophy, and cognitive decline in Alzheimer’s disease"

#### Tables


| Table no. | Title |
|---|---|
| **Table S1** | Demographic characteristics of the subsample with longitudinal data |
| **Table S2** | Associations between baseline tau PET or MRI and longitudinal cognition in CU&CI A+ |
| **Table S3** | Differences between baseline imaging predictors of longitudinal cognition in CU&CI A+ |
| **Table S4** | Associations between baseline tau PET or MRI and longitudinal cognition in CU A+ |
| **Table S5** | Associations between baseline tau PET or MRI and longitudinal cognition in CI A+ |
| **Table S6** | Associations between longitudinal tau PET or MRI and longitudinal cognition in CU&CI A+ |
| **Table S7** | Differences between longitudinal imaging predictors of longitudinal cognition in CU&CI A+ |
| **Table S8** | Associations between longitudinal tau PET or MRI and longitudinal cognition in CU A+ |
| **Table S9** | Associations between longitudinal tau PET or MRI and longitudinal cognition in CI A+ |
| **Table S10** | Associations between longitudinal tau PET or MRI and longitudinal cognition with and without baseline correction in CU&CI A+ |
| **Table S11** | Associations between longitudinal tau PET or MRI and longitudinal cognition with and without baseline correction in CU A+ |
| **Table S12** | Associations between longitudinal tau PET or MRI and longitudinal cognition with and without baseline correction in CI A+ |
| **Table S13** | Mediation effects of (longitudinal) tau pathology. (longitudinal) neurodegeneration. and longitudinal MMSE in CU&CI A+ |
| **Table S14** | Mediation effects of (longitudinal) tau pathology. (longitudinal) neurodegeneration. and longitudinal mPACC5 in CU&CI A+ |
| **Table S15a** | Cognitive slopes over 4 years based on simulated interventions on inferior middle temporal tau PET slopes in CU&CI A+ |
| **Table S15b** | Cognitive slopes over 4 years based on simulated interventions on neocortical tau PET slopes in CU&CI A+ |
| **Table S16a** | Differences in cognitive trajectories based on simulated reductions of inferior middle temporal tau PET slopes in CU&CI A+ |
| **Table S16b** | Differences in cognitive trajectories based on simulated reductions of neocortical tau PET slopes in CU&CI A+ |
| **Table S17a** | Differences between cognitive trajectories based on simulated reductions of inferior middle temporal tau PET slopes in CU A+ |
| **Table S17b** | Differences between cognitive trajectories based on simulated reductions of neocortical tau PET slopes in CU A+ |
| **Table S18a** | Differences between cognitive trajectories based on simulated reductions of inferior middle temporal tau PET slopes in CI A+ |
| **Table S18b** | Differences between cognitive trajectories based on simulated reductions of neocortical tau PET slopes in CI A+ |


#### Figures


| Figure no. | Title |
|---|---|
| **Figure S1** | Associations between baseline tau PET and MRI and longitudinal in CU&CI A+ showing inferior middle temporal |
| **Figure S2** | Associations between baseline tau PET and MRI and longitudinal cognition in CU A+ |
| **Figure S3** | Associations between baseline tau PET and MRI and longitudinal cognition in CI A+ |
| **Figure S4** | Associations between longitudinal tau PET and MRI and longitudinal cognition in CU A+ |
| **Figure S5** | Associations between longitudinal tau PET and MRI and longitudinal cognition in CI A+ |
| **Figure S6** | Reductions of effects of longitudinal tau PET and MRI vs longitudinal cognition in CU&CI A+ after baseline correction |
| **Figure S7** | Cognitive trajectories based on simulated reductions of tau PET slopes in CU A+ and CI A+ |

#### Table S1 Demographic characteristics of the subsample with longitudinal data

| | CU A+ (*n*=120) | CI A+ (*n*=202) | Overall (*n*=322) | p-value |
|---|---|---|---|---|
| Age, years | 70 (± 8.4) | 73 (± 7.4) | 72 (± 7.9) | 0.00462 |
| Female, *n* (%) | 64 (53 %) | 93 (46 %) | 157 (49 %) | 0.25 |
| Education, years | 13 (± 3.7) | 12 (± 4.4) | 13 (± 4.1) | 0.703 |
| MMSE | 29 (± 1.4) | 24 (± 4.6) | 26 (± 4.4) | <0.001 |
| mPACC5 | -0.25 (± 0.75) | -2.7 (± 1.7) | -1.8 (± 1.9) | <0.001 |
| Tau PET SUVR, entorhinal | 1.4 (± 0.30) | 1.8 (± 0.47) | 1.6 (± 0.46) | <0.001 |
| Tau PET SUVR, amygdala | 1.1 (± 0.32) | 1.5 (± 0.52) | 1.4 (± 0.51) | <0.001 |
| Tau PET SUVR, inferior middle temporal | 1.3 (± 0.30) | 1.9 (± 0.76) | 1.7 (± 0.68) | <0.001 |
| Tau PET SUVR, neocortical | 1.1 (± 0.12) | 1.3 (± 0.41) | 1.2 (± 0.36) | <0.001 |
| MRI cortical thickness, entorhinal | 3.2 (± 0.31) | 2.8 (± 0.40) | 3.0 (± 0.42) | <0.001 |
| MRI cortical volume, hippocampus | 0.0024 (± 0.00031) | 0.0020 (± 0.00034) | 0.0021 (± 0.00037) | <0.001 |
| MRI cortical thickness, inferior middle temporal | 2.5 (± 0.11) | 2.4 (± 0.14) | 2.4 (± 0.14) | <0.001 |
| MRI cortical thickness, neocortical | 2.3 (± 0.089) | 2.2 (± 0.11) | 2.3 (± 0.11) | <0.001 |
| MMSE slope | -0.29 (± 0.54) | -1.9 (± 1.3) | -1.3 (± 1.3) | <0.001 |
| mPACC5 slope | -0.10 (± 0.25) | -0.61 (± 0.40) | -0.40 (± 0.42) | <0.001 |
| Tau PET SUVR slope, entorhinal | 0.032 (± 0.047) | 0.044 (± 0.083) | 0.039 (± 0.072) | 0.155 |
| Tau PET SUVR slope, amygdala | 0.035 (± 0.047) | 0.062 (± 0.086) | 0.052 (± 0.075) | 0.00165 |
| Tau PET SUVR slope, inferior middle temporal | 0.033 (± 0.066) | 0.12 (± 0.13) | 0.085 (± 0.12) | <0.001 |
| Tau PET SUVR slope, neocortical | 0.014 (± 0.035) | 0.066 (± 0.083) | 0.047 (± 0.074) | <0.001 |
| MRI cortical thickness slope, entorhinal | -0.038 (± 0.049) | -0.073 (± 0.073) | -0.060 (± 0.067) | <0.001 |
| MRI cortical volume slope, hippocampus | -0.000042 (± 0.000034) | -0.000066 (± 0.000048) | -0.000057 (± 0.000045) | <0.001 |
| MRI cortical thickness slope, inferior middle temporal | -0.0078 (± 0.022) | -0.036 (± 0.037) | -0.026 (± 0.035) | <0.001 |
| MRI cortical thickness slope, neocortical | -0.0089 (± 0.014) | -0.020 (± 0.025) | -0.016 (± 0.022) | <0.001 |

Data are shown as mean (standard deviation) unless otherwise specified.

#### Table S2 Associations between baseline tau PET or MRI and longitudinal cognition in CU&CI A+

| Cogtest | Modality | Region | Estimate unadjusted | SE | p-value | p.FDR | Estimate adjusted | SE | p-value | p.FDR | %Reduction |
|---|---|---|---|---|---|---|---|---|---|---|---|
| MMSE | Tau | tau entorhinal | -0.1023 | 0.0136 | 0.000000 | 0.000000 | -0.0842 | 0.0139 | 0.000000 | 0.000000 | 17.69 |
| | | tau aymgdala | -0.0757 | 0.0139 | 0.000000 | 0.000000 | -0.0441 | 0.0139 | 0.001609 | 0.002574 | 41.74 |
| | | tau inf mid temporal | -0.1831 | 0.0128 | 0.000000 | 0.000000 | -0.1639 | 0.0142 | 0.000000 | 0.000000 | 10.49 |
| | | tau neocortical | -0.1680 | 0.0137 | 0.000000 | 0.000000 | -0.1588 | 0.0141 | 0.000000 | 0.000000 | 5.48 |
| | MRI | thickness entorhinal | -0.0891 | 0.0142 | 0.000000 | 0.000000 | -0.0626 | 0.0143 | 0.000017 | 0.000036 | 29.74 |
| | | volume hippocampus | -0.0828 | 0.0148 | 0.000000 | 0.000000 | -0.0693 | 0.0155 | 0.000010 | 0.000024 | 16.30 |
| | | thickness inf mid temporal | -0.1131 | 0.0131 | 0.000000 | 0.000000 | -0.0385 | 0.0130 | 0.003196 | 0.004511 | 65.96 |
| | | thickness neocortical | -0.0726 | 0.0138 | 0.000000 | 0.000000 | -0.0301 | 0.0129 | 0.019960 | 0.023951 | 58.54 |
| mPACC5 | Tau | tau entorhinal | -0.1146 | 0.0158 | 0.000000 | 0.000000 | -0.1024 | 0.0162 | 0.000000 | 0.000000 | 10.65 |
| | | tau aymgdala | -0.0812 | 0.0164 | 0.000001 | 0.000002 | -0.0589 | 0.0160 | 0.000272 | 0.000467 | 27.46 |
| | | tau inf mid temporal | -0.2223 | 0.0168 | 0.000000 | 0.000000 | -0.2142 | 0.0178 | 0.000000 | 0.000000 | 3.64 |
| | | tau neocortical | -0.2528 | 0.0168 | 0.000000 | 0.000000 | -0.2468 | 0.0170 | 0.000000 | 0.000000 | 2.37 |
| | MRI | thickness entorhinal | -0.0744 | 0.0168 | 0.000013 | 0.000016 | -0.0449 | 0.0166 | 0.007164 | 0.009049 | 39.65 |
| | | volume hippocampus | -0.0841 | 0.0172 | 0.000002 | 0.000002 | -0.0680 | 0.0176 | 0.000137 | 0.000253 | 19.14 |
| | | thickness inf mid temporal | -0.0943 | 0.0167 | 0.000000 | 0.000000 | -0.0195 | 0.0150 | 0.195968 | 0.213783 | 79.32 |
| | | thickness neocortical | -0.0655 | 0.0168 | 0.000118 | 0.000122 | -0.0229 | 0.0137 | 0.096233 | 0.109981 | 65.04 |

Data are shown as mean (standard deviation) unless otherwise specified.

#### Table S3 Differences between baseline imaging predictors of longitudinal cognition in CU&CI A+

| Test | Contrast | Difference | 95%CI|Lower, Upper] | p-value |
|---|---|---|---|---|
| MMSE | Tau PET inferior middle temporal vs Tau PET neocortical | -0.01241104 | -0.0233477656, -0.0002892816 | <0.05 |
| | Tau PET neocortical vs MRI inferior middle temporal | -0.2177035 | -0.2491835, -0.1873207 | <0.05 |
| | MRI inferior middle temporal vs Tau PET entorhinal | 0.1662452 | 0.1370352, 0.1962421 | <0.05 |
| | Tau PET entorhinal vs MRI entorhinal | -0.1718659 | -0.2034357, -0.1432902 | <0.05 |
| | MRI entorhinal vs MRI hippocampus | -0.01257386 | -0.028371160, 0.003466048 | ns |
| | MRI hippocampus vs MRI neocortical | 0.03953801 | 0.01955526, 0.06027265 | <0.05 |
| | MRI neocortical vs tau PET amygdala | 0.121862 | 0.09283711, 0.14889035 | <0.05 |
| mPACC5 | Tau PET neocortical vs Tau PET inferior middle temporal | -0.0183928 | -0.031432268, -0.005890647 | <0.05 |
| | Tau PET inferior middle temporal vs tau PET entorhinal | -0.07315993 | -0.08819253, -0.05798952 | <0.05 |
| | Tau PET entorhinal vs MRI inferior middle temporal | -0.1864504 | -0.2182821, -0.1569562 | <0.05 |
| | MRI inferior middle temporal vs MRI hippocampus | -0.004018812 | -0.02533052, 0.01706913 | ns |
| | MRI hippocampus vs tau PET amygdala | 0.1550845 | 0.1265288, 0.1866044 | <0.05 |
| | Tau PET amygdala vs MRI entorhinal | -0.1440105 | -0.1755606, -0.1154867 | <0.05 |
| | MRI entorhinal vs MRI neocortical | 0.01408771 | -0.004854687, 0.034243975 | ns |

Data are shown as mean (standard deviation) unless otherwise specified. *Note*: data were bootstrapped using n=1000 iterations.

#### Table S4 Associations between baseline tau PET or MRI and longitudinal cognition in CU A+

| Cogtest | Modality | Region | Estimate unadjusted | SE | P | p.FDR | Label | Estimate adjusted | SE | p-value | p.FDR | Label | %Reduction |
|---|---|---|---|---|---|---|---|---|---|---|---|---|---|
| MMSE | Tau | tau entorhinal | -0.0782 | 0.0197 | 0.0001 | 0.0002 | ** | -0.0650 | 0.0205 | 0.0019 | 0.0041 | ** | 16.9453 |
| | | tau aymgdala | -0.0809 | 0.0207 | 0.0001 | 0.0002 | ** | -0.0284 | 0.0221 | 0.2014 | 0.2302 | | 64.9513 |
| | | tau inf mid temporal | -0.2006 | 0.0236 | 0.0000 | 0.0000 | ** | -0.1817 | 0.0259 | 0.0000 | 0.0000 | ** | 9.4278 |
| | | tau neocortical | -0.2046 | 0.0329 | 0.0000 | 0.0000 | ** | -0.1903 | 0.0332 | 0.0000 | 0.0000 | ** | 6.9921 |
| | MRI | thickness entorhinal | -0.0564 | 0.0181 | 0.0023 | 0.0029 | ** | -0.0383 | 0.0184 | 0.0393 | 0.0589 | * | 32.0800 |
| | | volume hippocampus | -0.0606 | 0.0174 | 0.0007 | 0.0009 | ** | -0.0521 | 0.0185 | 0.0057 | 0.0101 | ** | 14.0682 |
| | | thickness inf mid temporal | -0.0730 | 0.0167 | 0.0000 | 0.0001 | ** | -0.0266 | 0.0156 | 0.0927 | 0.1271 | | 63.6286 |
| | | thickness neocortical | -0.0472 | 0.0166 | 0.0053 | 0.0062 | ** | -0.0294 | 0.0152 | 0.0556 | 0.0785 | | 37.6287 |
| mPACC5 | Tau | tau entorhinal | -0.1003 | 0.0204 | 0.0000 | 0.0000 | ** | -0.0782 | 0.0206 | 0.0002 | 0.0006 | ** | 22.0433 |
| | | tau aymgdala | -0.0865 | 0.0220 | 0.0001 | 0.0002 | ** | -0.0314 | 0.0223 | 0.1614 | 0.2013 | | 63.6589 |
| | | tau inf mid temporal | -0.2012 | 0.0288 | 0.0000 | 0.0000 | ** | -0.1929 | 0.0310 | 0.0000 | 0.0000 | ** | 4.1611 |
| | | tau neocortical | -0.1852 | 0.0391 | 0.0000 | 0.0000 | ** | -0.1726 | 0.0394 | 0.0000 | 0.0001 | ** | 6.8378 |
| | MRI | thickness entorhinal | -0.0865 | 0.0183 | 0.0000 | 0.0000 | ** | -0.0649 | 0.0183 | 0.0006 | 0.0015 | ** | 24.9839 |
| | | volume hippocampus | -0.0863 | 0.0177 | 0.0000 | 0.0000 | ** | -0.0770 | 0.0188 | 0.0001 | 0.0003 | ** | 10.7588 |
| | | thickness inf mid temporal | -0.0573 | 0.0197 | 0.0043 | 0.0052 | ** | -0.0134 | 0.0189 | 0.4810 | 0.5131 | | 76.6881 |
| | | thickness neocortical | -0.0431 | 0.0185 | 0.0216 | 0.0229 | ** | -0.0297 | 0.0178 | 0.0978 | 0.1304 | | 31.1986 |

Data are shown as mean (standard deviation) unless otherwise specified.

#### Table S5 Associations between baseline tau PET or MRI and longitudinal cognition in CI A+

| Cogtest | Modality | Region | Estimate unadjusted | SE | p-value | p.FDR | Label | Estimate adjusted | SE | p-value | p.FDR | Label | %Reduction |
|---|---|---|---|---|---|---|---|---|---|---|---|---|---|
| MMSE | Tau | tau entorhinal | -0.1085 | 0.0177 | 0.0000 | 0.0000 | ** | -0.0888 | 0.0182 | 0.0000 | 0.0000 | ** | 18.1745 |
| | | tau aymgdala | -0.0743 | 0.0179 | 0.0000 | 0.0001 | ** | -0.0471 | 0.0178 | 0.0085 | 0.0145 | ** | 36.6321 |
| | | tau inf mid temporal | -0.1853 | 0.0161 | 0.0000 | 0.0000 | ** | -0.1655 | 0.0180 | 0.0000 | 0.0000 | ** | 10.6819 |
| | | tau neocortical | -0.1767 | 0.0171 | 0.0000 | 0.0000 | ** | -0.1688 | 0.0180 | 0.0000 | 0.0000 | ** | 4.4731 |
| | MRI | thickness entorhinal | -0.0978 | 0.0188 | 0.0000 | 0.0000 | ** | -0.0687 | 0.0193 | 0.0004 | 0.0012 | ** | 29.7849 |
| | | volume hippocampus | -0.0891 | 0.0204 | 0.0000 | 0.0000 | ** | -0.0751 | 0.0213 | 0.0005 | 0.0013 | ** | 15.6848 |
| | | thickness inf mid temporal | -0.1245 | 0.0173 | 0.0000 | 0.0000 | ** | -0.0418 | 0.0178 | 0.0194 | 0.0300 | ** | 66.4163 |
| | | thickness neocortical | -0.0807 | 0.0186 | 0.0000 | 0.0000 | ** | -0.0251 | 0.0179 | 0.1636 | 0.2013 | | 68.9651 |
| mPACC5 | Tau | tau entorhinal | -0.1194 | 0.0216 | 0.0000 | 0.0000 | ** | -0.1107 | 0.0223 | 0.0000 | 0.0000 | ** | 7.3494 |
| | | tau aymgdala | -0.0796 | 0.0221 | 0.0004 | 0.0006 | ** | -0.0667 | 0.0215 | 0.0022 | 0.0045 | ** | 16.1977 |
| | | tau inf mid temporal | -0.2290 | 0.0219 | 0.0000 | 0.0000 | ** | -0.2206 | 0.0233 | 0.0000 | 0.0000 | ** | 3.6844 |
| | | tau neocortical | -0.2787 | 0.0211 | 0.0000 | 0.0000 | ** | -0.2748 | 0.0216 | 0.0000 | 0.0000 | ** | 1.4051 |
| | MRI | thickness entorhinal | -0.0655 | 0.0237 | 0.0063 | 0.0071 | ** | -0.0325 | 0.0237 | 0.1722 | 0.2066 | | 50.3908 |
| | | volume hippocampus | -0.0779 | 0.0251 | 0.0022 | 0.0028 | ** | -0.0605 | 0.0257 | 0.0193 | 0.0300 | ** | 22.3275 |
| | | thickness inf mid temporal | -0.1066 | 0.0232 | 0.0000 | 0.0000 | ** | -0.0220 | 0.0211 | 0.2992 | 0.3340 | | 79.3565 |
| | | thickness neocortical | -0.0746 | 0.0238 | 0.0020 | 0.0027 | ** | -0.0137 | 0.0190 | 0.4731 | 0.5131 | | 81.6794 |

Data are shown as mean (standard deviation) unless otherwise specified.

#### Table S6 Associations between longitudinal tau PET or MRI and longitudinal cognition in CU&CI A+

| Cogtest | Modality | Region | Estimate unadjusted | SE | p-value | p.FDR | Estimate adjusted | SE | p-value | p.FDR | %Reduction |
|---|---|---|---|---|---|---|---|---|---|---|---|
| MMSE | Tau | tau entorhinal | -0.3444 | 0.0483 | 0.000000 | 0.000000 | -0.2660 | 0.0500 | 0.000000 | 0.000000 | 22.76 |
| | | tau aymgdala | -0.2849 | 0.0501 | 0.000000 | 0.000000 | -0.1822 | 0.0473 | 0.000144 | 0.000189 | 36.05 |
| | | tau inf mid temporal | -0.4753 | 0.0451 | 0.000000 | 0.000000 | -0.2583 | 0.0463 | 0.000000 | 0.000000 | 45.66 |
| | | tau neocortical | -0.5606 | 0.0440 | 0.000000 | 0.000000 | -0.4562 | 0.0454 | 0.000000 | 0.000000 | 18.62 |
| | MRI | thickness entorhinal | -0.3199 | 0.0493 | 0.000000 | 0.000000 | -0.2264 | 0.0504 | 0.000010 | 0.000015 | 29.23 |
| | | volume hippocampus | -0.4184 | 0.0546 | 0.000000 | 0.000000 | -0.3646 | 0.0552 | 0.000000 | 0.000000 | 12.86 |
| | | thickness inf mid temporal | -0.5540 | 0.0415 | 0.000000 | 0.000000 | -0.4241 | 0.0459 | 0.000000 | 0.000000 | 23.45 |
| | | thickness neocortical | -0.4182 | 0.0452 | 0.000000 | 0.000000 | -0.2514 | 0.0426 | 0.000000 | 0.000000 | 39.89 |
| mPACC5 | Tau | tau entorhinal | -0.3187 | 0.0529 | 0.000000 | 0.000000 | -0.2534 | 0.0543 | 0.000005 | 0.000008 | 20.49 |
| | | tau aymgdala | -0.2292 | 0.0556 | 0.000049 | 0.000051 | -0.1397 | 0.0526 | 0.008412 | 0.008412 | 39.05 |
| | | tau inf mid temporal | -0.4699 | 0.0532 | 0.000000 | 0.000000 | -0.2824 | 0.0595 | 0.000003 | 0.000006 | 39.90 |
| | | tau neocortical | -0.6218 | 0.0521 | 0.000000 | 0.000000 | -0.5354 | 0.0562 | 0.000000 | 0.000000 | 13.90 |
| | MRI | thickness entorhinal | -0.2854 | 0.0530 | 0.000000 | 0.000000 | -0.2069 | 0.0538 | 0.000150 | 0.000189 | 27.51 |
| | | volume hippocampus | -0.3523 | 0.0589 | 0.000000 | 0.000000 | -0.3208 | 0.0594 | 0.000000 | 0.000000 | 8.94 |
| | | thickness inf mid temporal | -0.4860 | 0.0506 | 0.000000 | 0.000000 | -0.3387 | 0.0578 | 0.000000 | 0.000000 | 30.31 |
| | | thickness neocortical | -0.3767 | 0.0514 | 0.000000 | 0.000000 | -0.1797 | 0.0492 | 0.000307 | 0.000368 | 52.30 |

Data are shown as mean (standard deviation) unless otherwise specified.

#### Table S7 Differences between longitudinal imaging predictors of longitudinal cognition in CU&CI A+

| Test | Contrast | Difference | 95%CI[Lower, Upper] | p-value |
|---|---|---|---|---|
| MMSE | Tau PET neocortical vs MRI inferior middle temporal | -0.8026728 | -0.9784964, -0.5998986 | <0.05 |
| | MRI inferior middle temporal vs tau PET inferior middle temporal | 0.6711056 | 0.4921495, 0.8386747 | <0.05 |
| | Tau PET inferior middle temporal vs MRI neocortical | -0.4780032 | -0.6558373, -0.2921800 | <0.05 |
| | MRI neocortical vs MRI hippocampus | -0.02478903 | -0.1882316, 0.1326644 | ns |
| | MRI hippocampus vs Tau PET entorhinal | 0.2799341 | 0.1302846, 0.4263697 | <0.05 |
| | Tau PET entorhinal vs MRI entorhinal | -0.2718101 | -0.4166517, -0.1282120 | <0.05 |
| | MRI entorhinal vs tau PET amygdala | 0.2938822 | 0.1396872, 0.4388190 | <0.05 |
| mPACC5 | Tau neocortical vs MRI inferior middle temporal | -0.8982188 | -1.1372371, -0.6434743 | <0.05 |
| | MRI inferior middle temporal vs tau PET inferior middle temporal | 0.7191813 | 0.4797753, 0.9499966 | <0.05 |
| | Tau PET inferior middle temporal vs MRI neocortical | -0.5845741 | -0.8353325, -0.3301403 | <0.05 |
| | MRI neocortical vs MRI hippocampus | 0.08441033 | -0.09616297, 0.27506829 | ns |
| | MRI hippocampus vs tau PET entorhinal | 0.2032218 | 0.02386674, 0.37752609 | <0.05 |
| | Tau PET entorhinal vs MRI entorhinal | -0.2287765 | -0.39637370, -0.05311524 | <0.05 |
| | MRI entorhinal vs tau PET amygdala | 0.2625537 | 0.08080072, 0.45186680 | <0.05 |

Data are shown as mean (standard deviation) unless otherwise specified. *Note:* data were bootstrapped using n=1000 iterations.

#### Table S8 Associations between longitudinal tau PET or MRI and longitudinal cognition in CU A+

| Cogtest | Modality | Region | Estimate unadjusted | SE | p-value | p.FDR | Label | Estimate adjusted | SE | p-value | p.FDR | Label | %Reduction |
|---|---|---|---|---|---|---|---|---|---|---|---|---|---|
| MMSE | Tau | tau entorhinal | -0.1991 | 0.0572 | 0.0007 | 0.0009 | ** | -0.1643 | 0.0595 | 0.0068 | 0.0093 | ** | 17.4615 |
| | | tau aymgdala | -0.1420 | 0.0528 | 0.0083 | 0.0088 | ** | -0.0374 | 0.0546 | 0.4943 | 0.4943 | | 73.6336 |
| | | tau inf mid temporal | -0.3751 | 0.0639 | 0.0000 | 0.0000 | ** | -0.2172 | 0.0727 | 0.0035 | 0.0052 | ** | 42.1034 |
| | | tau neocortical | -0.5040 | 0.0844 | 0.0000 | 0.0000 | ** | -0.3558 | 0.0881 | 0.0001 | 0.0003 | ** | 29.4070 |
| | MRI | thickness entorhinal | -0.1332 | 0.0477 | 0.0062 | 0.0068 | ** | -0.0910 | 0.0489 | 0.0653 | 0.0712 | | 31.7306 |
| | | volume hippocampus | -0.1910 | 0.0517 | 0.0003 | 0.0004 | ** | -0.1806 | 0.0540 | 0.0011 | 0.0019 | ** | 5.4849 |
| | | thickness inf mid temporal | -0.3185 | 0.0492 | 0.0000 | 0.0000 | ** | -0.2224 | 0.0574 | 0.0002 | 0.0004 | ** | 30.1642 |
| | | thickness neocortical | -0.2660 | 0.0453 | 0.0000 | 0.0000 | ** | -0.1841 | 0.0471 | 0.0002 | 0.0004 | ** | 30.7794 |
| mPACC5 | Tau | tau entorhinal | -0.3174 | 0.0757 | 0.0001 | 0.0001 | ** | -0.2357 | 0.0761 | 0.0025 | 0.0038 | ** | 25.7392 |
| | | tau aymgdala | -0.2289 | 0.0705 | 0.0015 | 0.0018 | ** | -0.0931 | 0.0697 | 0.1844 | 0.1924 | | 59.3362 |
| | | tau inf mid temporal | -0.5363 | 0.0848 | 0.0000 | 0.0000 | ** | -0.4259 | 0.1011 | 0.0001 | 0.0002 | ** | 20.5900 |
| | | tau neocortical | -0.6654 | 0.1149 | 0.0000 | 0.0000 | ** | -0.5084 | 0.1232 | 0.0001 | 0.0003 | ** | 23.5906 |
| | MRI | thickness entorhinal | -0.2744 | 0.0615 | 0.0000 | 0.0000 | ** | -0.2138 | 0.0625 | 0.0009 | 0.0016 | ** | 22.0994 |
| | | volume hippocampus | -0.3481 | 0.0664 | 0.0000 | 0.0000 | ** | -0.3220 | 0.0689 | 0.0000 | 0.0001 | ** | 7.4853 |
| | | thickness inf mid temporal | -0.3440 | 0.0708 | 0.0000 | 0.0000 | ** | -0.1555 | 0.0798 | 0.0539 | 0.0601 | | 54.7803 |
| | | thickness neocortical | -0.3120 | 0.0635 | 0.0000 | 0.0000 | ** | -0.1950 | 0.0659 | 0.0037 | 0.0054 | ** | 37.4984 |

Data are shown as mean (standard deviation) unless otherwise specified.

#### Table S9 Associations between longitudinal tau PET or MRI and longitudinal cognition in CI A+

| Cogtest | Modality | Region | Estimate unadjusted | SE | p-value | P.FDR | Label | Estimate adjusted | SE | p-value | p.FDR | Label | %Reduction |
|---|---|---|---|---|---|---|---|---|---|---|---|---|---|
| MMSE | Tau | tau entorhinal | -0.3843 | 0.0661 | 0.0000 | 0.0000 | ** | -0.2857 | 0.0683 | 0.0000 | 0.0002 | ** | 25.6542 |
| | | tau aymgdala | -0.3277 | 0.0699 | 0.0000 | 0.0000 | ** | -0.2254 | 0.0641 | 0.0005 | 0.0011 | ** | 31.2337 |
| | | tau inf mid temporal | -0.5051 | 0.0604 | 0.0000 | 0.0000 | ** | -0.2711 | 0.0597 | 0.0000 | 0.0001 | ** | 46.3298 |
| | | tau neocortical | -0.6330 | 0.0586 | 0.0000 | 0.0000 | ** | -0.5264 | 0.0594 | 0.0000 | 0.0000 | ** | 16.8370 |
| | MRI | thickness entorhinal | -0.3916 | 0.0691 | 0.0000 | 0.0000 | ** | -0.2840 | 0.0711 | 0.0001 | 0.0003 | ** | 27.4695 |
| | | volume hippocampus | -0.5041 | 0.0787 | 0.0000 | 0.0000 | ** | -0.4408 | 0.0785 | 0.0000 | 0.0000 | ** | 12.5536 |
| | | thickness inf mid temporal | -0.6173 | 0.0553 | 0.0000 | 0.0000 | ** | -0.4842 | 0.0602 | 0.0000 | 0.0000 | ** | 21.5653 |
| | | thickness neocortical | -0.4656 | 0.0633 | 0.0000 | 0.0000 | ** | -0.2797 | 0.0572 | 0.0000 | 0.0000 | ** | 39.9253 |
| mPACC5 | Tau | tau entorhinal | -0.3131 | 0.0728 | 0.0000 | 0.0000 | ** | -0.2509 | 0.0753 | 0.0011 | 0.0019 | ** | 19.8810 |
| | | tau aymgdala | -0.2125 | 0.0789 | 0.0078 | 0.0084 | ** | -0.1464 | 0.0741 | 0.0498 | 0.0570 | * | 31.1134 |
| | | tau inf mid temporal | -0.4562 | 0.0723 | 0.0000 | 0.0000 | ** | -0.2496 | 0.0791 | 0.0019 | 0.0031 | ** | 45.2824 |
| | | tau neocortical | -0.6742 | 0.0698 | 0.0000 | 0.0000 | ** | -0.6007 | 0.0753 | 0.0000 | 0.0000 | ** | 10.9032 |
| | MRI | thickness entorhinal | -0.2842 | 0.0757 | 0.0002 | 0.0003 | ** | -0.2031 | 0.0773 | 0.0094 | 0.0126 | ** | 28.5302 |
| | | volume hippocampus | -0.3274 | 0.0870 | 0.0002 | 0.0003 | ** | -0.3009 | 0.0872 | 0.0007 | 0.0014 | ** | 8.0800 |
| | | thickness inf mid temporal | -0.5223 | 0.0689 | 0.0000 | 0.0000 | ** | -0.3924 | 0.0787 | 0.0000 | 0.0000 | ** | 24.8839 |
| | | thickness neocortical | -0.3839 | 0.0735 | 0.0000 | 0.0000 | ** | -0.1633 | 0.0680 | 0.0175 | 0.0221 | ** | 57.4556 |

Data are shown as mean (standard deviation) unless otherwise specified.

#### Table S10 Associations between longitudinal tau PET or MRI and cognitive decline with and without baseline correction in CU&CI A+

| Cogtest | Modality | Region | Estimate raw | SE | p-value | p.FDR | Label | Estimate corrected | SE | p-value | p.FDR | Label | %Reduction |
|---|---|---|---|---|---|---|---|---|---|---|---|---|---|
| MMSE | Tau | tau entorhinal | -0.3444 | 0.0483 | 0.0000 | 0.0000 | ** | 0.1399 | 0.1577 | 0.3758 | 0.4831 | | 140.62 |
| | | tau aymgdala | -0.2849 | 0.0501 | 0.0000 | 0.0000 | ** | 0.0220 | 0.0921 | 0.8112 | 0.8626 | | 107.73 |
| | | tau inf mid temporal | -0.4753 | 0.0451 | 0.0000 | 0.0000 | ** | 0.1935 | 0.0705 | 0.0064 | 0.0158 | ** | 140.70 |
| | | tau neocortical | -0.5606 | 0.0440 | 0.0000 | 0.0000 | ** | -0.0690 | 0.0977 | 0.4803 | 0.5895 | | 87.69 |
| | MRI | thickness entorhinal | -0.3199 | 0.0493 | 0.0000 | 0.0000 | ** | -0.1210 | 0.0630 | 0.0558 | 0.1158 | | 62.19 |
| | | volume hippocampus | -0.4184 | 0.0546 | 0.0000 | 0.0000 | ** | -0.9020 | 0.2676 | 0.0008 | 0.0025 | ** | -115.61 |
| | | thickness inf mid temporal | -0.5540 | 0.0415 | 0.0000 | 0.0000 | ** | -0.4251 | 0.0510 | 0.0000 | 0.0000 | ** | 23.27 |
| | | thickness neocortical | -0.4182 | 0.0452 | 0.0000 | 0.0000 | ** | -0.3047 | 0.0504 | 0.0000 | 0.0000 | ** | 27.13 |
| mPACC5 | Tau | tau entorhinal | -0.3187 | 0.0529 | 0.0000 | 0.0000 | ** | -0.0147 | 0.1713 | 0.9319 | 0.9319 | | 95.40 |
| | | tau aymgdala | -0.2292 | 0.0556 | 0.0000 | 0.0001 | ** | -0.0417 | 0.1015 | 0.6812 | 0.7997 | | 81.79 |
| | | tau inf mid temporal | -0.4699 | 0.0532 | 0.0000 | 0.0000 | ** | 0.0931 | 0.0893 | 0.2977 | 0.4230 | | 119.82 |
| | | tau neocortical | -0.6218 | 0.0521 | 0.0000 | 0.0000 | ** | -0.1221 | 0.1096 | 0.2664 | 0.3996 | | 80.37 |
| | MRI | thickness entorhinal | -0.2854 | 0.0530 | 0.0000 | 0.0000 | ** | -0.1675 | 0.0691 | 0.0160 | 0.0360 | ** | 41.32 |
| | | volume hippocampus | -0.3523 | 0.0589 | 0.0000 | 0.0000 | ** | -1.1974 | 0.2895 | 0.0000 | 0.0002 | ** | -239.83 |
| | | thickness inf mid temporal | -0.4860 | 0.0506 | 0.0000 | 0.0000 | ** | -0.4272 | 0.0616 | 0.0000 | 0.0000 | ** | 12.10 |
| | | thickness neocortical | -0.3767 | 0.0514 | 0.0000 | 0.0000 | ** | -0.3213 | 0.0575 | 0.0000 | 0.0000 | ** | 14.69 |

Data are shown as mean (standard deviation) unless otherwise specified.

#### Table S11 Associations between longitudinal tau PET or MRI and longitudinal cognition with and without baseline correction in CU A+

| Cogtest | Modality | Region | Estimate raw | SE | p-value | p.FDR | Label | Estimate corrected | SE | p-value | p.FDR | Label | %Reduction |
|---|---|---|---|---|---|---|---|---|---|---|---|---|---|
| MMSE | Tau | tau entorhinal | -0.1991 | 0.0572 | 0.0007 | 0.0009 | ** | -0.0385 | 0.1631 | 0.8139 | 0.8861 | | 80.67 |
| | | tau aymgdala | -0.1420 | 0.0528 | 0.0083 | 0.0088 | ** | -0.0457 | 0.0940 | 0.6277 | 0.7667 | | 67.79 |
| | | tau inf mid temporal | -0.3751 | 0.0639 | 0.0000 | 0.0000 | ** | 0.0891 | 0.1025 | 0.3865 | 0.5641 | | 123.75 |
| | | tau neocortical | -0.5040 | 0.0844 | 0.0000 | 0.0000 | ** | -0.2981 | 0.1301 | 0.0239 | 0.0585 | * | 40.86 |
| | MRI | thickness entorhinal | -0.1332 | 0.0477 | 0.0062 | 0.0068 | ** | -0.0357 | 0.0585 | 0.5435 | 0.7147 | | 73.24 |
| | | volume hippocampus | -0.1910 | 0.0517 | 0.0003 | 0.0004 | ** | -0.2507 | 0.2115 | 0.2384 | 0.4290 | | -31.20 |
| | | thickness inf mid temporal | -0.3185 | 0.0492 | 0.0000 | 0.0000 | ** | -0.2254 | 0.0581 | 0.0002 | 0.0011 | ** | 29.24 |
| | | thickness neocortical | -0.2660 | 0.0453 | 0.0000 | 0.0000 | ** | -0.1817 | 0.0528 | 0.0008 | 0.0040 | ** | 31.69 |
| mPACC5 | Tau | tau entorhinal | -0.3174 | 0.0757 | 0.0001 | 0.0001 | ** | -0.1016 | 0.2159 | 0.6389 | 0.7667 | | 68.00 |
| | | tau aymgdala | -0.2289 | 0.0705 | 0.0015 | 0.0018 | ** | -0.0739 | 0.1251 | 0.5559 | 0.7147 | | 67.73 |
| | | tau inf mid temporal | -0.5363 | 0.0848 | 0.0000 | 0.0000 | ** | -0.3583 | 0.1517 | 0.0199 | 0.0511 | * | 33.19 |
| | | tau neocortical | -0.6654 | 0.1149 | 0.0000 | 0.0000 | ** | -0.7377 | 0.1804 | 0.0001 | 0.0006 | ** | -10.86 |
| | MRI | thickness entorhinal | -0.2744 | 0.0615 | 0.0000 | 0.0000 | ** | -0.1273 | 0.0745 | 0.0901 | 0.1976 | | 53.60 |
| | | volume hippocampus | -0.3481 | 0.0664 | 0.0000 | 0.0000 | ** | -0.6396 | 0.2701 | 0.0196 | 0.0511 | * | -83.75 |
| | | thickness inf mid temporal | -0.3440 | 0.0708 | 0.0000 | 0.0000 | ** | -0.2875 | 0.0861 | 0.0011 | 0.0049 | ** | 16.40 |
| | | thickness neocortical | -0.3120 | 0.0635 | 0.0000 | 0.0000 | ** | -0.2298 | 0.0754 | 0.0029 | 0.0097 | ** | 26.36 |

Data are shown as mean (standard deviation) unless otherwise specified.

#### Table S12 Associations between longitudinal tau PET or MRI and longitudinal cognition with and without baseline correction in CI A+

| Cogtest | Modality | Region | Estimate raw | SE | p-value | p.FDR | Label | Estimate corrected | SE | p-value | p.FDR | Label | %Reduction |
|---|---|---|---|---|---|---|---|---|---|---|---|---|---|
| MMSE | Tau | tau entorhinal | -0.3843 | 0.0661 | 0.0000 | 0.0000 | ** | 0.1838 | 0.2219 | 0.4086 | 0.5807 | | 147.82 |
| | | tau aymgdala | -0.3277 | 0.0699 | 0.0000 | 0.0000 | ** | 0.0162 | 0.1301 | 0.9010 | 0.9297 | | 104.95 |
| | | tau hippocampus | -0.3079 | 0.0673 | 0.0000 | 0.0000 | ** | -0.0532 | 0.1470 | 0.7180 | 0.8249 | | 82.73 |
| | | tau inf mid temporal | -0.5051 | 0.0604 | 0.0000 | 0.0000 | ** | 0.1936 | 0.0922 | 0.0372 | 0.0873 | * | 138.33 |
| | | tau neocortical | -0.6330 | 0.0586 | 0.0000 | 0.0000 | ** | -0.0792 | 0.1281 | 0.5371 | 0.7147 | | 87.48 |
| | MRI | thickness entorhinal | -0.3916 | 0.0691 | 0.0000 | 0.0000 | ** | -0.1487 | 0.0917 | 0.1066 | 0.2132 | | 62.03 |
| | | volume hippocampus | -0.5041 | 0.0787 | 0.0000 | 0.0000 | ** | -1.4661 | 0.4277 | 0.0007 | 0.0040 | ** | -190.83 |
| | | thickness inf mid temporal | -0.6173 | 0.0553 | 0.0000 | 0.0000 | ** | -0.4739 | 0.0685 | 0.0000 | 0.0000 | ** | 23.24 |
| | | thickness neocortical | -0.4656 | 0.0633 | 0.0000 | 0.0000 | ** | -0.3369 | 0.0699 | 0.0000 | 0.0001 | ** | 27.64 |
| mPACC5 | Tau | tau entorhinal | -0.3131 | 0.0728 | 0.0000 | 0.0000 | ** | 0.0552 | 0.2427 | 0.8205 | 0.8861 | | 117.62 |
| | | tau aymgdala | -0.2125 | 0.0789 | 0.0078 | 0.0084 | ** | -0.0159 | 0.1448 | 0.9125 | 0.9297 | | 92.50 |
| | | tau hippocampus | -0.1856 | 0.0757 | 0.0153 | 0.0156 | ** | 0.0186 | 0.1558 | 0.9052 | 0.9297 | | 110.01 |
| | | tau inf mid temporal | -0.4562 | 0.0723 | 0.0000 | 0.0000 | ** | 0.1919 | 0.1143 | 0.0952 | 0.1976 | | 142.07 |
| | | tau neocortical | -0.6742 | 0.0698 | 0.0000 | 0.0000 | ** | 0.0121 | 0.1398 | 0.9311 | 0.9311 | | 101.80 |
| | MRI | thickness entorhinal | -0.2842 | 0.0757 | 0.0002 | 0.0003 | ** | -0.1734 | 0.1029 | 0.0940 | 0.1976 | | 39.00 |
| | | volume hippocampus | -0.3274 | 0.0870 | 0.0002 | 0.0003 | ** | -1.6139 | 0.4881 | 0.0012 | 0.0049 | ** | -392.95 |
| | | thickness inf mid temporal | -0.5223 | 0.0689 | 0.0000 | 0.0000 | ** | -0.4575 | 0.0841 | 0.0000 | 0.0000 | ** | 12.41 |
| | | thickness neocortical | -0.3839 | 0.0735 | 0.0000 | 0.0000 | ** | -0.3270 | 0.0810 | 0.0001 | 0.0006 | ** | 14.83 |

Data are shown as mean (standard deviation) unless otherwise specified.

**Table S13 Mediation effects of (longitudinal) tau pathology, (longitudinal) neurodegeneration, and longitudinal MMSE in CU&CI A+**

| Region | Path | Pathname | Estimate | CI lower | CI upper | p-value | Prop. mediated |
|---|---|---|---|---|---|---|---|
| Inf mid temporal | M1~X | a1 | -0.458 | -0.577 | 0.350 | <0.001 | |
| | M2~X | a2 | 0.880 | 0.771 | 0.985 | 0.985 | |
| | M3~X | a3 | -0.261 | -0.487 | 0.006 | 0.030 | |
| | M2~M2 | a4 | 0.104 | 0.034 | 0.188 | 0.008 | |
| | M3~M2 | d21 | 0.420 | 0.296 | 0.522 | <0.001 | |
| | Y~X | c'=direct | -0.481 | -0.709 | -0.255 | <0.001 | 0.90 |
| | Y~M1 | b1 | 0.021 | -0.097 | 0.139 | 0.727 | |
| | Y~M2 | b2 | 0.146 | -0.059 | 0.351 | 0.161 | |
| | Y~M3 | b3 | 0.278 | 0.155 | 0.411 | <0.001 | |
| | Indirect 1 | a3*b3 | -0.073 | -0.168 | -0.008 | * | 0.14 |
| | Indirect 2 | a1*d21*b3 | -0.054 | -0.092 | -0.029 | * | 0.10 |
| | Total | See below | -0.532 | -0.652 | -0.400 | * | |
| Neocortical | M1~X | a1 | -0.306 | -0.418 | 0.187 | <0.001 | |
| | M2~X | a2 | 0.892 | 0.823 | 0.962 | <0.001 | |
| | M3~X | a3 | -0.007 | -0.258 | 0.332 | 0.961 | |
| | M2~M2 | a4 | 0.037 | -0.014 | 0.108 | 0.221 | |
| | M3~M2 | d21 | 0.420 | 0.296 | 0.522 | <0.001 | |
| | Y~X | c'=direct | -0.260 | -0.532 | -0.008 | 0.050 | 0.54 |
| | Y~M1 | b1 | 0.112 | -0.012 | 0.248 | 0.091 | |
| | Y~M2 | b2 | -0.129 | -0.425 | 0.179 | 0.405 | |
| | Y~M3 | b3 | 0.181 | 0.077 | 0.306 | 0.002 | |
| | Indirect 1 | a2*b4*b3 | -0.049 | -0.120 | -0.010 | * | 0.10 |
| | Indirect 2 | a1*d21*b3 | -0.023 | -0.052 | -0.009 | * | 0.05 |
| | Total | See below | -0.481 | -0.616 | -0.369 | * | |

Data are shown as mean (standard deviation) unless otherwise specified.
*Note*: all analyses are corrected for age, sex, education and cognitive status.
Total=c'+(a1*b1)+(a2*b2)+(a3*b3)+(a1*a4*b2)+(a2*b4*b3)+(a1*d21*b3)+(a1*a4*b4*b3).
X=predictor (baseline tau PET), Y=outcome (longitudinal cognition)
M1=mediator 1 (baseline MRI), M2=mediatior 2 (longitudinal MRI), M3=mediator 3 (longitudinal tau PET)

#### Table S14 Mediation effects of (longitudinal) tau pathology, (longitudinal) neurodegeneration, and longitudinal mPACC5 in CU&CI A+

| Region | Path | Pathname | Estimate | CI lower | CI upper | p-value | Prop. mediated |
|---|---|---|---|---|---|---|---|
| Inf mid temporal | M1~X | a1 | -0.413 | -0.558 | -0.273 | <0.001 | |
| | M2~X | a2 | 0.873 | 0.756 | 0.988 | <0.001 | |
| | M3~X | a3 | -0.299 | -0.574 | -0.020 | 0.032 | |
| | M2~M2 | a4 | 0.112 | 0.029 | 0.193 | 0.008 | |
| | M3~M2 | d21 | 0.384 | 0.291 | 0.478 | <0.001 | |
| | Y~X | c'=direct | -0.472 | -0.754 | -0.156 | 0.001 | 0.82 |
| | Y~M1 | b1 | 0.012 | -0.129 | 0.144 | 0.865 | |
| | Y~M2 | b2 | 0.158 | -0.107 | 0.404 | 0.226 | |
| | Y~M3 | b3 | 0.251 | 0.099 | 0.435 | 0.004 | |
| | Indirect 1 | a3*b3 | -0.075 | -0.188 | -0.013 | * | 0.15 |
| | Indirect 2 | a1*d21*b3 | -0.040 | -0.076 | 0.017 | * | 0.08 |
| | Total | *See notes* | -0.501 | -0.670 | -0.336 | * | |
| Neocortical | M1~X | a1 | -0.224 | -0.353 | 0.111 | <0.001 | |
| | M2~X | a2 | 0.876 | 0.793 | 0.960 | <0.001 | |
| | M3~X | a3 | 0.042 | -0.203 | 0.374 | 0.770 | |
| | M2~M2 | a4 | 0.044 | -0.021 | 0.123 | 0.237 | |
| | M3~M2 | d21 | 0.401 | 0.274 | 0.509 | <0.001 | |
| | Y~X | c'=direct | -0.465 | -0.762 | -0.075 | 0.007 | 0.94 |
| | Y~M1 | b1 | 0.079 | -0.033 | 0.192 | 0.169 | |
| | Y~M2 | b2 | -0.022 | -0.323 | 0.250 | 0.881 | |
| | Y~M3 | b3 | 0.145 | 0.040 | 0.300 | 0.026 | |
| | Indirect 1 | a2*b4*b3 | -0.053 | -0.122 | -0.014 | * | 0.09 |
| | Indirect 2 | a1*d21*b3 | -0.013 | -0.040 | 0.003 | * | 0.02 |
| | Total | *See notes* | -0.561 | -0.731 | -0.378 | * | |

Data are shown as mean (standard deviation) unless otherwise specified.
*Note*: all analyses are corrected for age, sex, education and cognitive status.
Total=c'+(a1*b1)+(a2*b2)+(a3*b3)+(a1*a4*b2)+(a2*b4*b3)+(a1*d21*b3)+(a1*a4*b4*b3).
X=predictor (baseline tau PET), Y=outcome (longitudinal cognition)
M1=mediator 1 (baseline MRI), M2=mediatior 2 (longitudinal MRI), M3=mediator 3 (longitudinal tau PET)

#### Table S15a Cognitive slopes over 4 years based on simulated reductions of inferior middle temporal tau PET slopes in CU&CI A+

| | | CU A+ (n=116) | CI A+ (n=155) | Overall (n=271) | p-value |
|---|---|---|---|---|---|
| **MMSE** | **Placebo** | | | | |
| | Mean (SD) | -0.29 (± 0.40) | -1.9 (± 0.65) | -1.3 (± 0.99) | <0.001 |
| | **30% reduction** | | | | |
| | Mean (SD) | -0.23 (± 0.33) | -1.7 (± 0.46) | -1.2 (± 0.84) | <0.001 |
| | **50% reduction** | | | | |
| | Mean (SD) | -0.19 (± 0.28) | -1.6 (± 0.34) | -1.0 (± 0.75) | <0.001 |
| | **70% reduction** | | | | |
| | Mean (SD) | -0.15 (± 0.25) | -1.4 (± 0.24) | -0.95 (± 0.67) | <0.001 |
| | **No progression** | | | | |
| | Mean (SD) | 0.70 (± 0.15) | -0.96 (± 0.14) | -0.32 (± 0.82) | <0.001 |
| **mPACC5** | **Placebo** | | | | |
| | Mean (SD) | -0.097 (± 0.14) | -0.61 (± 0.19) | -0.39 (± 0.31) | <0.001 |
| | **30% reduction** | | | | |
| | Mean (SD) | -0.079 (± 0.12) | -0.55 (± 0.14) | -0.35 (± 0.27) | <0.001 |
| | **50% reduction** | | | | |
| | Mean (SD) | -0.067 (± 0.11) | -0.51 (± 0.10) | -0.32 (± 0.24) | <0.001 |
| | **70% reduction** | | | | |
| | Mean (SD) | -0.055 (± 0.097) | -0.47 (± 0.081) | -0.29 (± 0.22) | <0.001 |
| | **No progression** | | | | |
| | Mean (SD) | 0.21 (± 0.057) | -0.31 (± 0.050) | -0.086 (± 0.26) | <0.001 |

Data are shown as mean (standard deviation) unless otherwise specified.

#### Table S15b Cognitive slopes over 4 years based on simulated reductions of neocortical tau PET slopes in CU&CI A+

| | | CU A+ (n=116) | CI A+ (n=157) | Overall (n=273) | p-value |
|---|---|---|---|---|---|
| **MMSE** | **Placebo** | | | | |
| | Mean (SD) | -0.29 (± 0.46) | -1.9 (± 0.74) | -1.3 (± 1.0) | <0.001 |
| | **30% reduction** | | | | |
| | Mean (SD) | -0.24 (± 0.40) | -1.7 (± 0.51) | -1.2 (± 0.86) | <0.001 |
| | **50% reduction** | | | | |
| | Mean (SD) | -0.21 (± 0.37) | -1.6 (± 0.37) | -1.1 (± 0.76) | <0.001 |
| | **70% reduction** | | | | |
| | Mean (SD) | -0.18 (± 0.35) | -1.4 (± 0.28) | -0.95 (± 0.68) | <0.001 |
| | **No progression** | | | | |
| | Mean (SD) | 0.75 (± 0.15) | -0.91 (± 0.14) | -0.28 (± 0.82) | <0.001 |
| **mPACC5** | **Placebo** | | | | |
| | Mean (SD) | -0.097 (± 0.17) | -0.61 (± 0.23) | -0.39 (± 0.33) | <0.001 |
| | **30% reduction** | | | | |
| | Mean (SD) | -0.082 (± 0.16) | -0.55 (± 0.16) | -0.35 (± 0.28) | <0.001 |
| | **50% reduction** | | | | |
| | Mean (SD) | -0.072 (± 0.15) | -0.50 (± 0.12) | -0.32 (± 0.25) | <0.001 |
| | **70% reduction** | | | | |
| | Mean (SD) | -0.062 (± 0.14) | -0.46 (± 0.10) | -0.29 (± 0.23) | <0.001 |
| | **No progression** | | | | |
| | Mean (SD) | 0.23 (± 0.057) | -0.28 (± 0.050) | -0.063 (± 0.26) | <0.001 |

Data are shown as mean (standard deviation) unless otherwise specified.

#### Table S16a Differences in cognitive trajectories based on simulated reductions of inferior middle temporal tau PET slopes in CU&CI A+

| Region | Cognitive test | Contrast | Difference | CI lower | CI upper | p-value | p-Bonferroni |
|---|---|---|---|---|---|---|---|
| Inf mid temporal | MMSE | 30-placebo | 0.1566935 | -0.02393456 | 0.3373215 | 0.1243768 | ns |
| | | 50-placebo | 0.2611558 | 0.08052775 | 0.4417838 | 0.0007818 | ns |
| | | 70-placebo | 0.3656181 | 0.18499007 | 0.5462461 | 0.0000004 | * |
| | | No progression-placebo | 0.9877832 | 0.80715517 | 1.1684112 | 0.0000000 | * |
| | | 50-30 | 0.1044623 | -0.07616572 | 0.2850904 | 0.5108581 | ns |
| | | 70-30 | 0.2089246 | 0.02829659 | 0.3895527 | 0.0139559 | ns |
| | | 70-50 | 0.1044623 | -0.07616572 | 0.2850904 | 0.5108581 | ns |
| | | 30-no progression | -0.8310897 | -1.01171777 | -0.6504617 | 0.0000000 | * |
| | | 50-no progression | -0.7266274 | -0.90725546 | -0.5459994 | 0.0000000 | * |
| | | 70-no progression | -0.6221651 | -0.80279314 | -0.4415371 | 0.0000000 | * |
| | mPACC5 | 30-placebo | 0.04348138 | -0.018031219 | 0.10499398 | 0.3014050 | ns |
| | | 50-placebo | 0.07246896 | 0.010956367 | 0.13398156 | 0.0115393 | ns |
| | | 70-placebo | 0.10145655 | 0.039943953 | 0.16296915 | 0.0000705 | * |
| | | No progression-placebo | 0.30726356 | 0.245750966 | 0.36877616 | 0.0000000 | * |
| | | 50-30 | 0.02898759 | -0.032525012 | 0.09050018 | 0.6991990 | ns |
| | | 70-30 | 0.05797517 | -0.003537426 | 0.11948777 | 0.0757050 | ns |
| | | 70-50 | 0.02898759 | -0.032525012 | 0.09050018 | 0.6991990 | ns |
| | | 30-no progression | -0.26378219 | -0.325294783 | -0.20226959 | 0.0000000 | * |
| | | 50-no progression | -0.23479460 | -0.296307197 | -0.17328200 | 0.0000000 | * |
| | | 70-no progression | -0.20580701 | -0.267319611 | -0.14429442 | 0.0000000 | * |

Data are shown as mean (standard deviation) unless otherwise specified.

### Table S16b Differences in cognitive trajectories based on simulated reductions of neocortical tau PET slopes in CU&CI A+

| Region | Cognitive test | Contrast | Difference | CI lower | CI upper | p-value | p-Bonferroni |
|---|---|---|---|---|---|---|---|
| Neocortical | MMSE | 30-placebo | 0.1518844 | -0.02222313 | 0.3259919 | 0.1120786 | ns |
| | | 50-placebo | 0.2531407 | 0.07903313 | 0.4272482 | 0.0011012 | * |
| | | 70-placebo | 0.3543969 | 0.18028939 | 0.5285045 | 0.0000011 | * |
| | | No progression-placebo | 1.0338870 | 0.84865055 | 1.2191235 | 0.0000000 | * |
| | | 50-30 | 0.1012563 | -0.07285127 | 0.2753638 | 0.4401232 | ns |
| | | 70-30 | 0.2025125 | 0.02840500 | 0.3766201 | 0.0149709 | ns |
| | | 70-50 | 0.1012563 | -0.07285127 | 0.2753638 | 0.4401232 | ns |
| | | 30-no progression | -0.8831852 | -1.06842167 | -0.6979487 | 0.0000000 | * |
| | | 50-no progression | -0.7827173 | -0.96795376 | -0.5974808 | 0.0000000 | * |
| | | 70-no progression | -0.6822493 | -0.86748584 | -0.4970128 | 0.0000000 | * |
| | mPACC5 | 30-placebo | 0.04433277 | -0.016164177 | 0.10482971 | 0.2348273 | ns |
| | | 50-placebo | 0.07388794 | 0.013391001 | 0.13438489 | 0.0093098 | ns |
| | | 70-placebo | 0.10344312 | 0.042946178 | 0.16394007 | 0.0000702 | * |
| | | No progression-placebo | 0.33027951 | 0.266173029 | 0.39438599 | 0.0000000 | * |
| | | 50-30 | 0.02955518 | -0.030941766 | 0.09005212 | 0.5905978 | ns |
| | | 70-30 | 0.05911036 | -0.001386588 | 0.11960730 | 0.0583158 | ns |
| | | 70-50 | 0.02955518 | -0.030941766 | 0.09005212 | 0.5905978 | ns |
| | | 30-no progression | -0.28634289 | -0.350449369 | -0.22223640 | 0.0000000 | * |
| | | 50-no progression | -0.25705180 | -0.321158286 | -0.19294532 | 0.0000000 | * |
| | | 70-no progression | -0.22776072 | -0.291867202 | -0.16365424 | 0.0000000 | * |

Data are shown as mean (standard deviation) unless otherwise specified.

#### Table S17a Differences in cognitive trajectories based on simulated reductions of inferior middle temporal tau PET slopes in CU A+

| Region | Cognitive test | Contrast | Difference | CI lower | CI upper | p-value | p-Bonferroni |
|---|---|---|---|---|---|---|---|
| Inf mid temporal | MMSE | 30-placebo | 0.05815316 | -0.046986717 | 0.1632930 | 0.5540966 | ns |
| | | 50-placebo | 0.09692193 | -0.008217944 | 0.2020618 | 0.0870657 | ns |
| | | 70-placebo | 0.13569070 | 0.030550828 | 0.2408306 | 0.0040620 | * |
| | | No progression-placebo | 0.98778321 | 0.882643336 | 1.0929231 | 0.0000000 | * |
| | | 50-30 | 0.03876877 | -0.066371103 | 0.1439086 | 0.8512609 | ns |
| | | 70-30 | 0.07753754 | -0.027602331 | 0.1826774 | 0.2585984 | ns |
| | | 70-50 | 0.03876877 | -0.066371103 | 0.1439086 | 0.8512609 | ns |
| | | 30-no progression | -0.92963005 | -1.034769927 | -0.8244902 | 0.0000000 | * |
| | | 50-no progression | -0.89086128 | -0.996001155 | -0.7857214 | 0.0000000 | * |
| | | 70-no progression | -0.85209251 | -0.957232383 | -0.7469526 | 0.0000000 | * |
| | mPACC5 | 30-placebo | 0.01795972 | -0.020680051 | 0.05659949 | 0.7086249 | ns |
| | | 50-placebo | 0.02993286 | -0.008706905 | 0.06857263 | 0.2129801 | ns |
| | | 70-placebo | 0.04190601 | 0.003266240 | 0.08054578 | 0.0258680 | ns |
| | | No progression-placebo | 0.30726356 | 0.268623795 | 0.34590333 | 0.0000000 | * |
| | | 50-30 | 0.01197315 | -0.026666624 | 0.05061291 | 0.9153954 | ns |
| | | 70-30 | 0.02394629 | -0.014693478 | 0.06258606 | 0.4372824 | ns |
| | | 70-50 | 0.01197315 | -0.026666624 | 0.05061291 | 0.9153954 | ns |
| | | 30-no progression | -0.28930385 | -0.327943615 | -0.25066408 | 0.0000000 | * |
| | | 50-no progression | -0.27733070 | -0.315970469 | -0.23869093 | 0.0000000 | * |
| | | 70-no progression | -0.26535755 | -0.303997324 | -0.22671779 | 0.0000000 | * |

Data are shown as mean (standard deviation) unless otherwise specified.

#### Table S17b Differences in cognitive trajectories based on simulated reductions of neocortical tau PET slopes in CU A+

| Region | Cognitive test | Contrast | Difference | CI lower | CI upper | p-value | p-Bonferroni |
|---|---|---|---|---|---|---|---|
| Neocortical | MMSE | 30-placebo | 0.03952507 | -0.03659901 | 0.1156492 | 0.5387081 | ns |
| | | 50-placebo | 0.06587512 | -0.01024897 | 0.1419992 | 0.1163721 | ns |
| | | 70-placebo | 0.09222516 | 0.01610108 | 0.1683492 | 0.1988727 | ns |
| | | No progression-placebo | 1.03388704 | 0.90462730 | 1.1631468 | 0.0000000 | * |
| | | 50-30 | 0.02635005 | -0.04977404 | 0.1024741 | 0.8087825 | ns |
| | | 70-30 | 0.05270009 | -0.02342399 | 0.1288242 | 0.2818588 | ns |
| | | 70-50 | 0.02635005 | -0.04977404 | 0.1024741 | 0.8087825 | ns |
| | | 30-no progression | -0.99027552 | -1.11953526 | -0.8610158 | 0.0000000 | * |
| | | 50-no progression | -0.96120116 | -1.09046091 | -0.8319414 | 0.0000000 | * |
| | | 70-no progression | -0.93212681 | -1.06138656 | -0.8028671 | 0.0000000 | * |
| | mPACC5 | 30-placebo | 0.01675792 | -0.025349578 | 0.05886542 | 0.7341882 | ns |
| | | 50-placebo | 0.02792987 | -0.014177630 | 0.07003737 | 0.3194429 | ns |
| | | 70-placebo | 0.03910182 | -0.003005682 | 0.08120932 | 0.0795583 | ns |
| | | No progression-placebo | 0.33027951 | 0.28025475 | 0.38030427 | 0.0000000 | * |
| | | 50-30 | 0.01117195 | -0.030935552 | 0.05327945 | 0.9031350 | ns |
| | | 70-30 | 0.02234390 | -0.019763604 | 0.06445140 | 0.5199860 | ns |
| | | 70-50 | 0.01117195 | -0.030935552 | 0.05327945 | 0.9031350 | ns |
| | | 30-no progression | -0.31525126 | -0.36527603 | -0.26522650 | 0.0000000 | * |
| | | 50-no progression | -0.30523243 | -0.35525719 | -0.25520767 | 0.0000000 | * |
| | | 70-no progression | -0.29521360 | -0.34523836 | -0.24518884 | 0.0000000 | * |

Data are shown as mean (standard deviation) unless otherwise specified.

#### Table S18a Differences in cognitive trajectories based on simulated reductions of inferior middle temporal tau PET slopes in CI A+

| Region | Cognitive test | Contrast | Difference | CI lower | CI upper | p-value | p-Bonferroni |
|---|---|---|---|---|---|---|---|
| Inf mid temporal | MMSE | 30-placebo | 0.2178922 | 0.10418030 | 0.3316041 | 0.0000020 | * |
| | | 50-placebo | 0.3631537 | 0.24944177 | 0.4768656 | 0.0000000 | * |
| | | 70-placebo | 0.5084151 | 0.39470323 | 0.6221270 | 0.0000000 | * |
| | | No progression-placebo | 0.9877832 | 0.87407132 | 1.1014951 | 0.0000000 | * |
| | | 50-30 | 0.1452615 | 0.03154957 | 0.2589734 | 0.0045677 | * |
| | | 70-30 | 0.2905229 | 0.17681104 | 0.4042348 | 0.0000000 | * |
| | | 70-50 | 0.1452615 | 0.03154957 | 0.2589734 | 0.0045677 | * |
| | | 30-no progression | -0.7698910 | -0.88360291 | -0.6561791 | 0.0000000 | * |
| | | 50-no progression | -0.6246295 | -0.73834144 | -0.5109177 | 0.0000000 | * |
| | | 70-no progression | -0.4793681 | -0.59307998 | -0.3656562 | 0.0000000 | * |
| | mPACC5 | 30-placebo | 0.06258146 | 0.024913302 | 0.10024962 | 0.0000631 | * |
| | | 50-placebo | 0.10430243 | 0.066634276 | 0.14197059 | 0.0000000 | * |
| | | 70-placebo | 0.14602341 | 0.108355250 | 0.18369157 | 0.0000000 | * |
| | | No progression-placebo | 0.30726356 | 0.269595406 | 0.34493172 | 0.0000000 | * |
| | | 50-30 | 0.04172097 | 0.004052815 | 0.07938913 | 0.0213462 | ns |
| | | 70-30 | 0.08344195 | 0.045773789 | 0.12111011 | 0.0000000 | * |
| | | 70-50 | 0.04172097 | 0.004052815 | 0.07938913 | 0.0213462 | ns |
| | | 30-no progression | -0.24468210 | -0.282350262 | -0.20701395 | 0.0000000 | * |
| | | 50-no progression | -0.20296113 | -0.240629288 | -0.16529297 | 0.0000000 | * |
| | | 70-no progression | -0.16124016 | -0.198908314 | -0.12357200 | 0.0000000 | * |

Data are shown as mean (standard deviation) unless otherwise specified.

#### Table S18b Differences in cognitive trajectories based on simulated reductions of neocortical tau PET slopes in CI A+

| Region | Cognitive test | Contrast | Difference | CI lower | CI upper | p-value | p-Bonferroni |
|---|---|---|---|---|---|---|---|
| Neocortical | MMSE | Placebo-30 | 0.2463959 | 0.08715943 | 0.4056324 | 0.0004308 | * |
| | | Placebo-50 | 0.4106599 | 0.25142338 | 0.5698964 | 0.0000000 | * |
| | | Placebo-70 | 0.5749238 | 0.41568733 | 0.7341603 | 0.0000000 | * |
| | | Placebo-no progression | 1.0338870 | 0.90604290 | 1.1617312 | 0.0000000 | * |
| | | 50-30 | 0.1642640 | 0.00502745 | 0.3235005 | 0.0402095 | ns |
| | | 70-30 | 0.3285279 | 0.16929140 | 0.4877644 | 0.0000008 | * |
| | | 70-50 | 0.1642640 | 0.00502745 | 0.3235005 | 0.0402095 | ns |
| | | 30-no progression | -0.8166764 | -0.94452058 | -0.6888323 | 0.0000000 | * |
| | | 50-no progression | -0.6718694 | -0.79971350 | -0.5440252 | 0.0000000 | * |
| | | 70-no progression | -0.5270623 | -0.65490643 | -0.3992181 | 0.0000000 | * |
| | mPACC5 | Placebo-30 | 0.07240030 | 0.016973192 | 0.1278274 | 0.0045065 | * |
| | | Placebo-50 | 0.12066717 | 0.065240060 | 0.1760943 | 0.0000002 | * |
| | | Placebo-70 | 0.16893404 | 0.113506929 | 0.2243611 | 0.0000000 | * |
| | | Placebo-no progression | 0.33027951 | 0.284803010 | 0.37575601 | 0.0000000 | * |
| | | 50-30 | 0.04826687 | -0.007160242 | 0.1036940 | 0.1128890 | ns |
| | | 70-30 | 0.09653374 | 0.041106626 | 0.1519608 | 0.0000509 | * |
| | | 70-50 | 0.04826687 | -0.007160242 | 0.1036940 | 0.1128890 | ns |
| | | 30-no progression | -0.26470823 | -0.310184732 | -0.21923173 | 0.0000000 | * |
| | | 50-no progression | -0.22099404 | -0.266470544 | -0.17551754 | 0.0000000 | * |
| | | 70-no progression | -0.17727985 | -0.222756356 | -0.13180335 | 0.0000000 | * |

Data are shown as mean (standard deviation) unless otherwise specified.

**Figure S1 Associations between baseline tau PET and MRI and longitudinal cognition in CU&CI A+ showing inferior middle temporal**

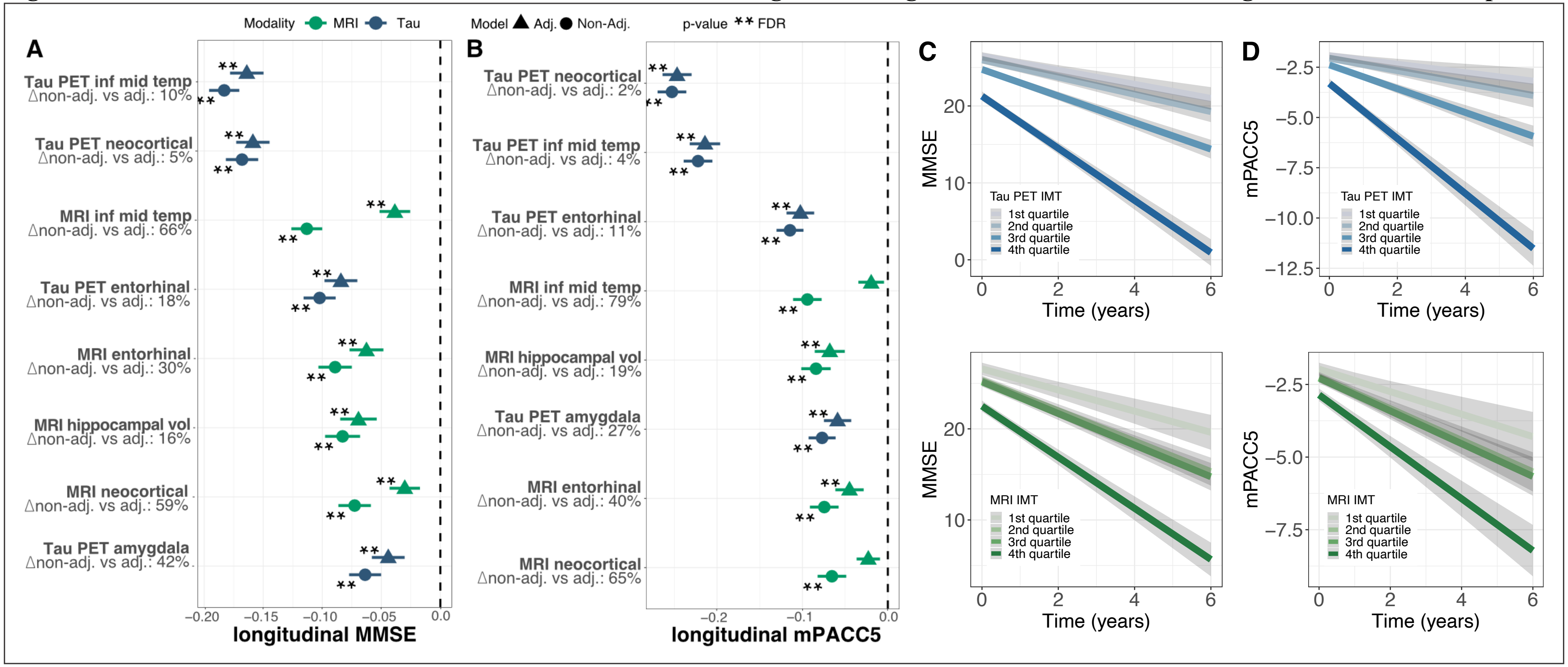


**(A)** shows associations between baseline tau PET and MRI and MMSE in the whole cohort (CU Aβ+ and CI Aβ+ individuals combined) while adjusting for age, sex, education and cognitive status (CU vs CI). **(B)** shows these associations for the mPACC5. Standardized coefficients for tau PET are depicted in blue, and standardized coefficients for MRI are depicted in green. Circles represent models without adjustment for the other modality, while triangles represent models with adjustment for the other imaging modality. **=significant after FDR correction, *=significant at $p$=0.05. **(C)** and **(D)** show changes in MMSE and mPACC5 by quartiles of neocortical tau PET (blue) or MRI (green), adjusted for age, sex and education.

**Figure S2 Associations between baseline tau PET and MRI and longitudinal cognition in CU A+**

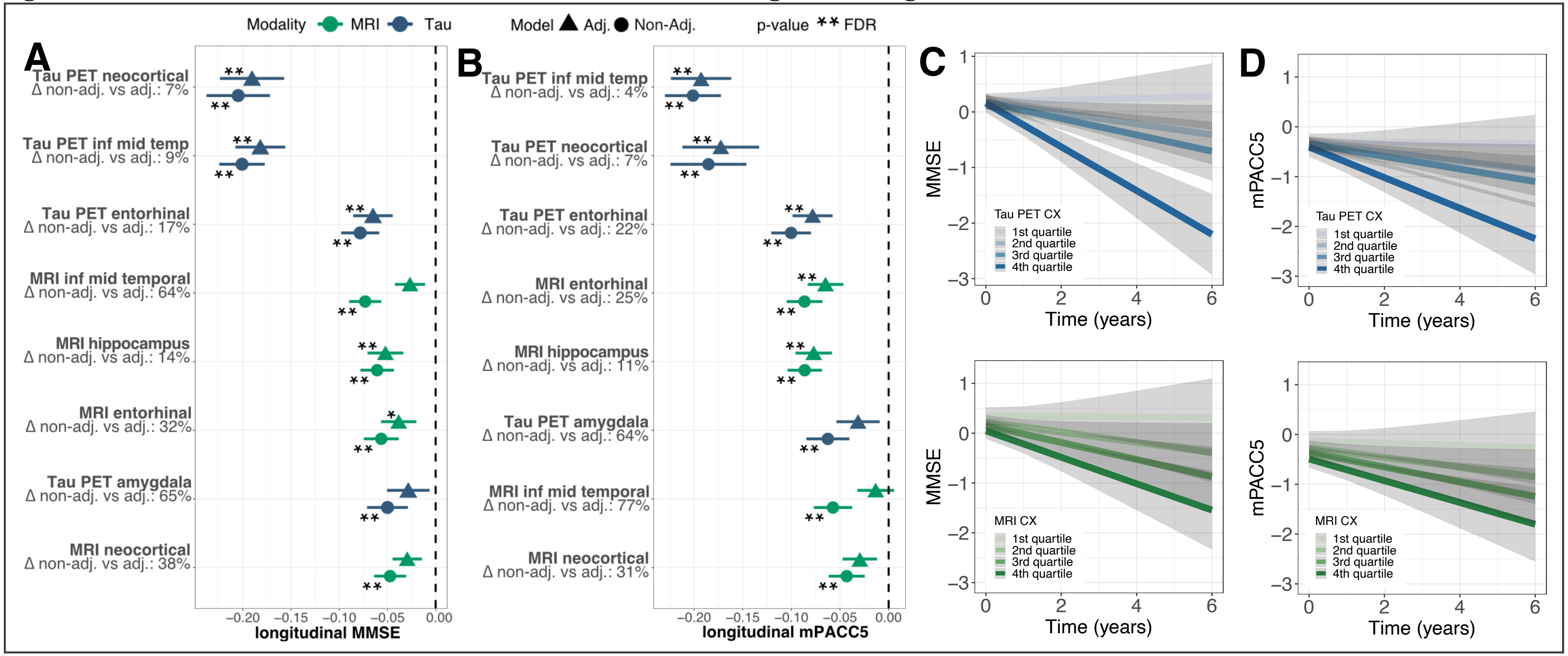


**(A)** shows associations between baseline tau PET and MRI and MMSE in CU Aβ+ individuals while adjusting for age, sex, education. **(B)** shows these associations for the mPACC5. Standardized coefficients for tau PET are depicted in blue, and standardized coefficients for MRI are depicted in green. Circles represent models without adjustment for the other modality, while triangles represent models with adjustment for the other imaging modality. **=significant after FDR correction, *=significant at $p$=0.05. **(C)** and **(D)** show changes in MMSE and mPACC5 by quartiles of neocortical tau PET (blue) or MRI (green), adjusted for age, sex and education.

**Figure S3 Associations between baseline tau PET and MRI and longitudinal cognition in CI A+**

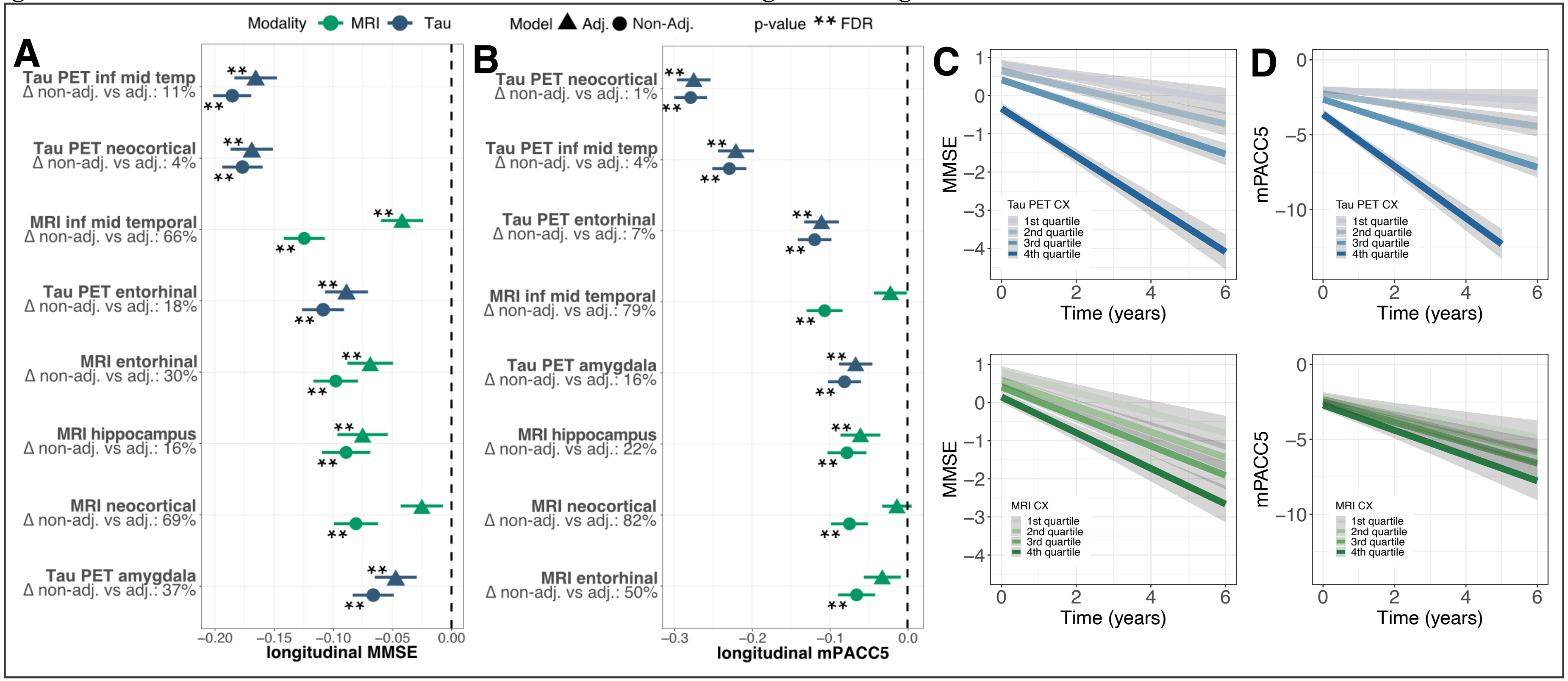


**(A)** shows associations between baseline tau PET and MRI and MMSE in CI Aβ+ individuals while adjusting for age, sex, education. **(B)** shows these associations for the mPACC5. Standardized coefficients for tau PET are depicted in blue, and standardized coefficients for MRI are depicted in green. Circles represent models without adjustment for the other modality, while triangles represent models with adjustment for the other imaging modality. **=significant after FDR correction, *=significant at $p$=0.05. **(C)** and **(D)** show changes in MMSE and mPACC5 by quartiles of neocortical tau PET (blue) or MRI (green), adjusted for age, sex and education.

**Figure S4 Associations between longitudinal tau PET and MRI and longitudinal cognition in CU A+**

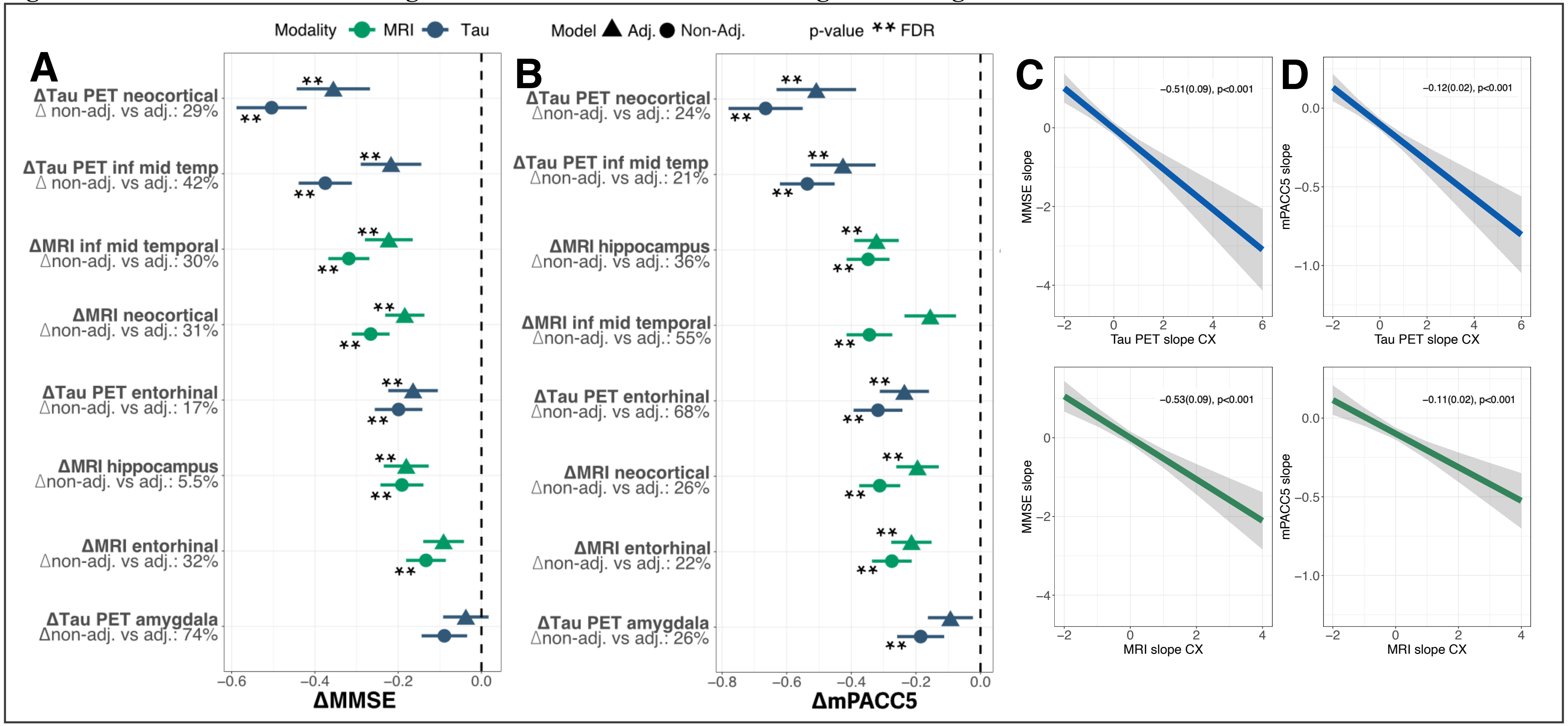


**(A)** shows associations between slope in tau PET and MRI and slope in MMSE in CU Aβ+ individuals while adjusting for age, sex, education. **(B)** shows these associations for the mPACC5. The slopes were obtained from linear mixed models with random slopes and random intercepts, assessing either tau~time, mri~time, or cognition~time, yielding average rate of change metrics per individual. These were used as input for the linear regressions depicted in these panels. **(C)** and **(D)** show associations between rate of change in MMSE and mPACC5 and rate of change in tau PET or MRI in regions of interest. **=significant after FDR correction, *=significant at p=0.05.

**Figure S5 Associations between longitudinal tau PET and MRI and longitudinal cognition in CI A+**

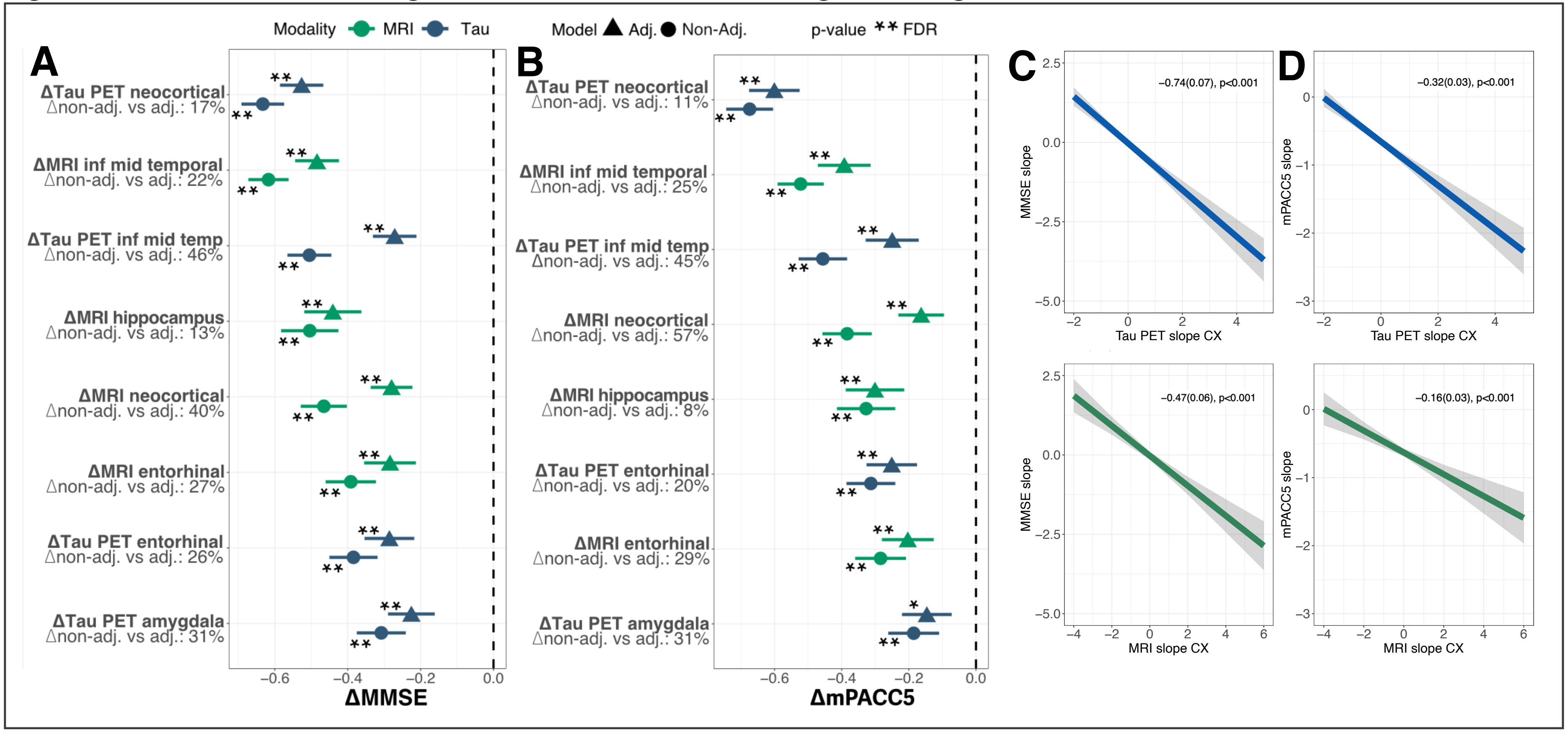


**(A)** shows associations between slope in tau PET and MRI and slope in MMSE in CI Aβ+ individuals while adjusting for age, sex, education. **(B)** shows these associations for the mPACC5. The slopes were obtained from linear mixed models with random slopes and random intercepts, assessing either tau~time, mri~time, or cognition~time, yielding average rate of change metrics per individual. These were used as input for the linear regressions depicted in these panels. **(C)** and **(D)** show associations between rate of change in MMSE and mPACC5 and rate of change in tau PET or MRI in regions of interest. **=significant after FDR correction, *=significant at p=0.05.

**Figure S6 Reductions of effects of longitudinal tau PET and MRI vs longitudinal cognition in CU&CI A+ after baseline correction**

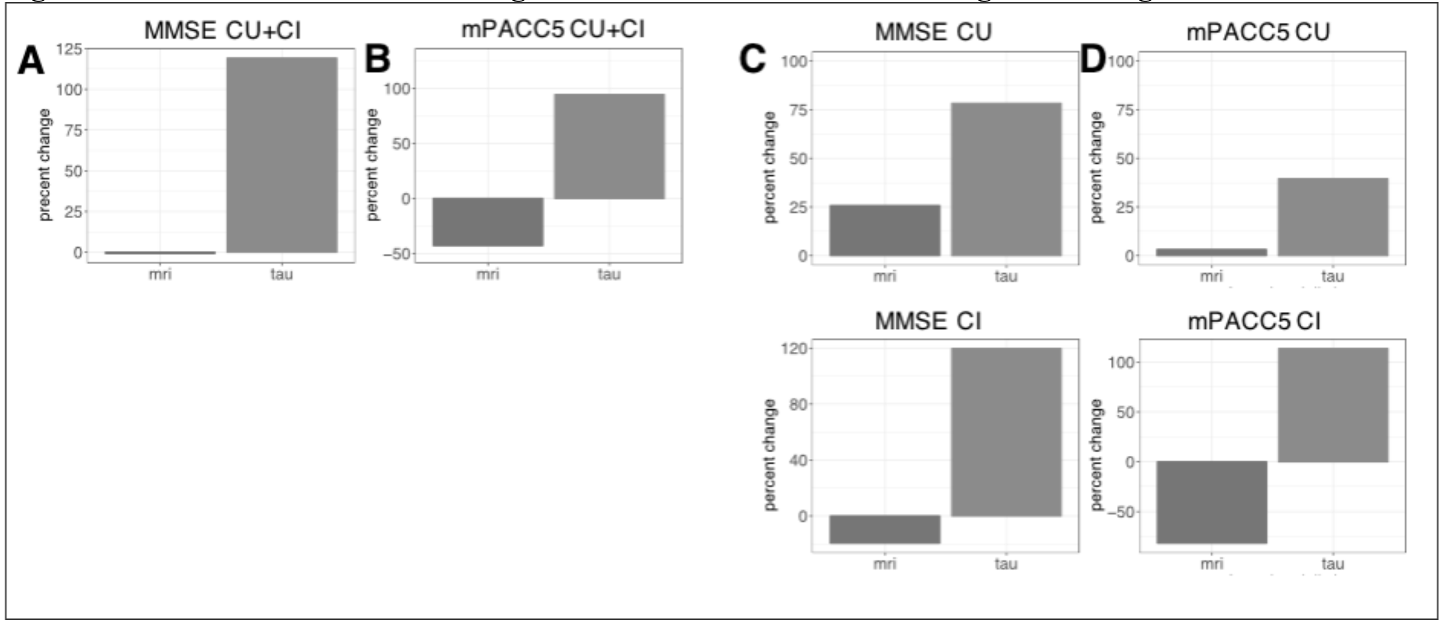


**(A)**and **(B)** show percentages change for tau PET or MRI based on correction for baseline levels of the respective imaging modalities for MMSE and mPACC5 respectively, while **(C)** and **(D)** show these percentages change for CU A+ and CI A+ individuals separately.

**Figure S7 Cognitive trajectories based on simulated reductions of tau PET slopes in CU A+ and CI A+**

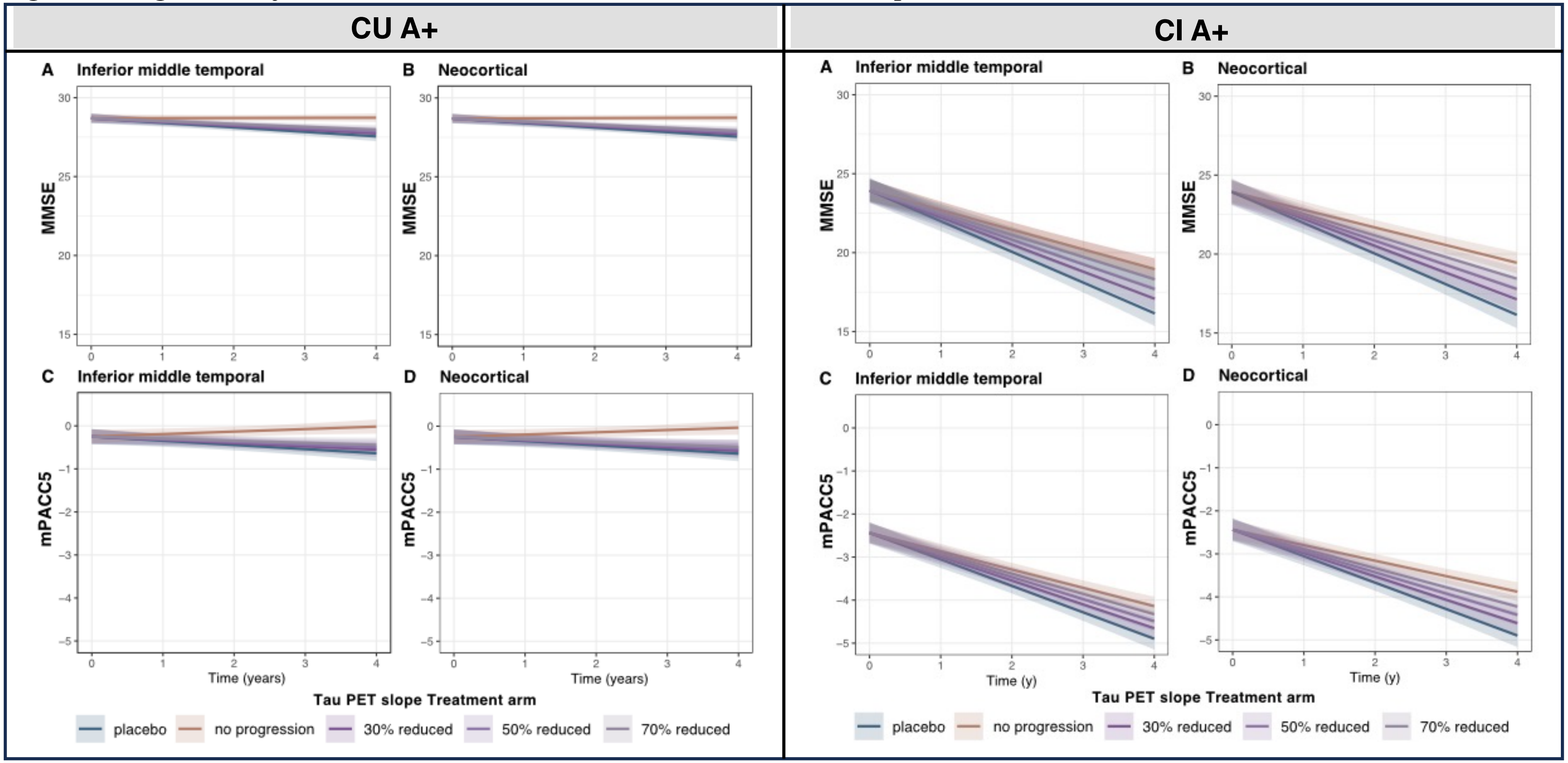


**Left (A)** and **(B)** show predicted MMSE values over time in inferior middle temporal and neocortical regions respectively in CU Aβ+ individuals in different arms of tau PET slopes, i.e. placebo (no intervention) versus 30%, 50% and 70% reductions of tau PET slopes, while **(C)** and **(D)** show predicted mPACC5 values over time in inferior middle temporal and neocortical regions respectively in the whole cohort (CU Aβ+ and CI Aβ+ individuals combined). **Right (A)**, **(B)**, **(C)**, **(D)** show the same for CI Aβ+ individuals.
